# Personalized phosphoproteomics of skeletal muscle insulin resistance and exercise links MINDY1 to insulin action

**DOI:** 10.1101/2024.03.11.24304084

**Authors:** Elise J. Needham, Janne R. Hingst, Johan D. Onslev, Alexis Diaz-Vegas, Magnus R. Leandersson, Jonas Møller Kristensen, Kohei Kido, Erik A. Richter, Kurt Højlund, Benjamin L. Parker, Kristen Cooke, Guang Yang, Christian Pehmøller, Sean J. Humphrey, David E. James, Jørgen F.P. Wojtaszewski

## Abstract

Type 2 diabetes is preceded by a defective insulin response, yet our knowledge of the precise mechanisms is incomplete. Here, we investigate how insulin resistance alters signalling responses in skeletal muscle and how this is modified by exercise. We measured parallel phenotypes and phosphoproteomes of insulin resistant and insulin sensitive individuals as they responded to exercise and insulin (n=19, 114 biopsies), quantifying over 12,000 phosphopeptides in each biopsy. Our personalized phosphoproteomics approach revealed that insulin resistant individuals have selective and time-dependent signalling alterations. Insulin resistant subjects have reduced insulin-stimulated mTORC1 responses and alterations to non-canonical rather than canonical insulin signalling. Prior exercise promotes insulin sensitivity even in insulin resistant individuals by ‘priming’ a portion of insulin signalling prior to insulin infusion. This includes MINDY1 S441, which is elevated in insulin-sensitive subjects and primed by prior exercise. MINDY1 contains a missense variant that is protective for type 2 diabetes but its role in disease risk is unknown. We show that MINDY1 S441 phosphorylation is downstream of AKT, and MINDY1 knockdown enhances insulin-stimulated glucose uptake in rat myotubes. This work delineates the signalling alterations in insulin resistant skeletal muscle and how exercise partially counteracts these and identifies MINDY1 as a regulator of insulin action.

**Highlights:**

- Insulin resistance primarily alters non-canonical insulin signalling.
- mTORC1 substrates were most defective in insulin resistance.
- Exercise counteracts insulin signalling defects including MINDY1 S441.
- MINDY1 is a negative regulator of insulin sensitivity in rat myotubes and S441 is downstream of AKT.

## Introduction

Protein phosphorylation facilitates rapid responses to stimuli to control cellular outcomes^1,2^. Insulin activates a large and complex network of protein phosphorylation events to maintain homeostasis^3,4^. Environmental factors like diet and physical activity as well as genetics play a key role in the emergence of insulin resistance, a state of impaired regulation of glucose metabolism by insulin. These effects can be rapid, as a single bout of exercise can increase skeletal muscle insulin sensitivity for up to 48 hours^5^. Insulin resistance occurs primarily in skeletal muscle, liver, and adipose tissue^6^, and is one of the earliest defects associated with a suite of metabolic diseases including type 2 diabetes (T2D) and cardiovascular disease (CVD). Rare, monogenic forms of severe insulin resistance involve defects in core insulin signalling proteins, such as INSR, IRS, and PI3K^7,8^. However, in the more common non-monogenic forms of insulin resistance, studies have identified no consistent defects in these proximal signalling components^9–11^. Global and unbiased measures of insulin signalling in insulin sensitive and resistant individuals are therefore needed to pinpoint sites of defective signalling. Studies in mice and human cell lines originally derived from insulin-resistant individuals have revealed extensive differential signalling between insulin sensitive and resistant organisms^10,12–14^. To reveal which of these signalling changes are most relevant to the effects of insulin resistance on glucose uptake in humans, we concurrently measured global signalling and glucose uptake, directly in skeletal muscle of humans with a broad range of insulin sensitivity under conditions that dynamically perturb insulin signalling.

Recent advances in mass spectrometry (MS)-based phosphoproteomics have enabled widespread quantification of phosphorylation dynamics in diverse biological contexts and organisms^15–18^. Global studies in skeletal muscle, adipose tissue, and liver of rodents and humans have revealed that insulin regulates thousands of phosphosites on hundreds of proteins^3,4,19–23^. A major challenge now lies in identifying the connectivity and functionally annotating these signalling networks. The extent of this challenge is highlighted by the observation that in these studies only ∼10% of proteins with phosphosites regulated by insulin belong to the “canonical” insulin signalling pathway. This is compounded by the unknown upstream regulation and downstream function of ∼95% of reported human phosphorylation sites^24^. We previously developed an experimental and computational framework called “personalized phosphoproteomics” that identifies signalling linked with a particular phenotype of interest^19^. By measuring phosphoproteomes and phenotypes in parallel, together with their dynamic responses to perturbations affecting each, this approach enables the contextualisation of global signalling responses.

Here, we quantified the skeletal muscle phosphoproteomes of insulin resistant and insulin sensitive individuals responding to insulin and exercise. We sampled six skeletal muscle biopsies from each individual after they recovered from a one-legged exercise bout for four hours, followed by a hyperinsulinemic-euglycemic clamp with sampling at 0, 30, and 120 minutes of the clamp. This experimental design enables the comparison of the effects of prior exercise and insulin resistance on the acute response to insulin, and with temporal resolution. By applying recent developments in sample preparation methods^4,25^ together with data independent acquisition (DIA) mass spectrometry^26^, we reliably quantified >26,000 phosphopeptides in human skeletal muscle. We extended our personalized phosphoproteomics framework by introducing a temporal component to further disentangle upstream versus responsive signalling. By integrating our phosphoproteome findings with candidates from published glycaemic trait genome-wide association studies, we prioritised potential causal mediators from the uncharacterized signalling we found to be defective in insulin resistant skeletal muscle. Missense variants in MINDY1 are protective for type 2 diabetes, but the cell types and mechanisms by which MINDY1 mediates these effects are unknown. We linked MINDY1 to the insulin signalling network by identifying AKT as an upstream kinase and found that MINDY1 is a negative regulator of insulin-stimulated glucose uptake in rat myotubes. Our definition and prioritisation of signalling alterations in insulin resistance and exercise-mediated insulin sensitization reveal selective and time-dependent alterations.

## Results

### Defining insulin sensitivity in lean and overweight individuals

We designed an experimental workflow to probe the metabolic and molecular differences between insulin resistant (IR) and insulin sensitive (IS) skeletal muscle, and how this is influenced by prior exercise. Healthy, untrained (VO_2peak_ 3.1±0.5 L min^-1^) male subjects aged 29±4 years (n=19) were recruited and segregated into groups of lean and IS in the fasted state (HOMA-IR < 1.5, BMI < 25 kg/m^2^, n = 10) and overweight and IR in the fasted state (HOMA-IR >= 2.2, BMI = 28-35 kg/m^2^, n = 9) (**Figure 1A, Table S1**). All subjects were glucose tolerant, indicating normal insulin secretory function (**Table S1**). To measure the subjects’ phenotypic and phosphoproteomic responses to exercise and insulin, subjects underwent 1h of single-legged knee-extensor exercise. After a 4h recovery, we biopsied the *vastus lateralis* muscle from exercised and non-exercised legs. We subsequently performed a 120 min hyperinsulinemic-euglycemic clamp, during which we biopsied each leg at 30 and 120 min.

**Figure 1:**
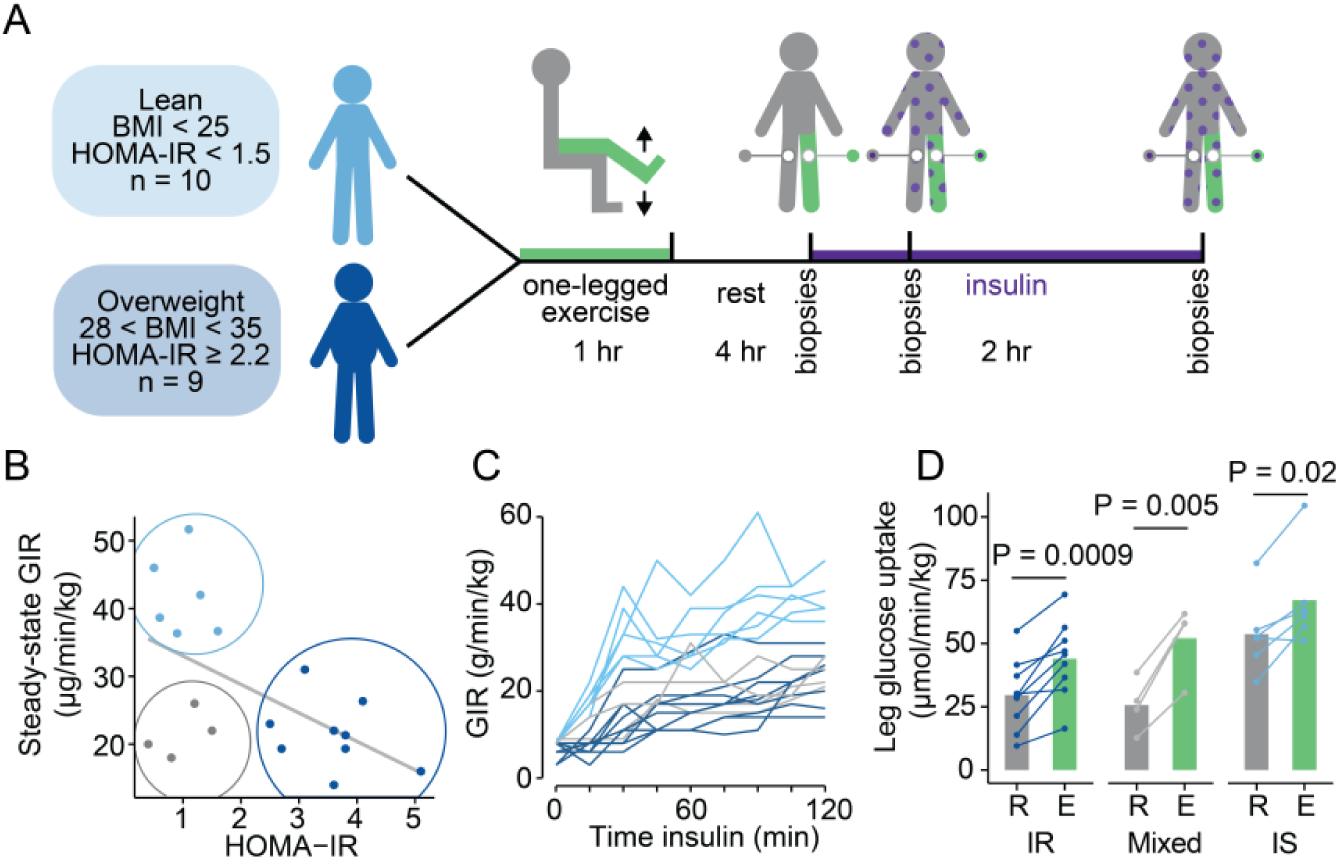
Defining insulin sensitivity by HOMA-IR, GIR and leg glucose uptake. **(A)** Study design depicting human subjects undergoing one-legged exercise followed by a hyperinsulinemic-euglycemic clamp. **(B)** HOMA-IR and steady state whole body glucose infusion rate (GIR) during the last 30 minutes of the hyperinsulinemic-euglycemic clamp. **(C)** Whole body GIR during the hyperinsulinemic euglycemic clamp. **(D)** Leg-specific glucose uptake into the rested (R) and exercised (E) legs of the subjects, separated into insulin sensitive (IS), insulin resistant (IR) groups, and a mixed HOMA-IR insulin sensitive, leg glucose uptake insulin resistant group (“mix”).

Phenotypic diversity was evident even between individuals who were lean and insulin sensitive. Four of the ten individuals deemed insulin sensitive based on HOMA-IR values were IR based on their glucose infusion rate (GIR) during the clamp (**Figure 1B**), consistent with previous studies^27,28^. We therefore based our classification on clamp data rather than HOMA-IR throughout our study. GIR varied by 3.4-fold across all individuals (**Figure 1C**). As expected, we did not observe differences in insulin-mediated suppression of hepatic glucose output (*P* = 0.15), as we used a relatively high insulin concentration^29,30^. Since our study is principally concerned with muscle insulin sensitivity and signalling, we separated this group of HOMA-IR insulin sensitive but GIR insulin resistant individuals (n=4, “mixed”) from the lean insulin sensitive group (n=6, “IS”) for further comparisons with the insulin resistant (n=9, “IR”) group (**Figure 1B**). Thus, under our study conditions, variation in GIR principally reflects changes in insulin regulation of whole-body glucose utilisation (Rd). As muscle makes a major contribution to Rd, we measured the amount of glucose taken up into rested and exercised legs in each individual during the clamp. We calculated the insulin-stimulated steady-state leg glucose uptake (LGU) for each individual as the average of the 90 min, 105 min and 120 min values during the insulin clamp (**Table S2**). IS individuals had on average 1.9-fold higher LGU during the hyperinsulinemic euglycemic clamp (*P* = 0.0006). This did not appear to be due to changes in bulk blood flow (**Figure S1A**). Prior exercise potentiated the LGU during the insulin clamp in both IS and IR individuals (IS *P* = 0.02, IR *P* = 0.0009) (**Figure 1D**). Insulin-stimulated LGU in the exercised leg of IR subjects approached the levels of the rested leg in IS subjects. Exercise sensitisation in IR individuals has previously been reported at the whole-body level^31^, and our study now demonstrates that this effect occurs directly at the level of skeletal muscle in IR subjects. Prior exercise can therefore partially counteract the phenotypic effects of skeletal muscle insulin resistance.

### Total Proteome

To determine if insulin resistance involves alterations in the levels of signalling components, we measured skeletal muscle proteomes of biopsies from the non-exercised legs of the subjects immediately prior to the insulin clamp (0 min). Employing offline high-pH reversed-phase chromatography and high-resolution data independent acquisition (DIA) mass spectrometry we quantified 5,905 proteins in total. Quantification coverage and reproducibility of our proteomics measurements was very high, with 96% of these proteins quantified in at least half of the subjects (**Figure 2A, Figure S2A, Table S3**), and a mean correlation co-efficient of 0.98 between individuals (**Figure S2B**). Dynamic range of measured protein intensities spanned seven orders of magnitude, with highly abundant proteins enriched in biological processes including muscle filament sliding, electron transport and the tricarboxylic acid cycle (**Figure 2B, Table S4**). Skeletal muscle proteomes separated by subject GIR in principal component 4 in PCA (**Figure S2C**). To determine how insulin resistance rewires skeletal muscle, we performed differential expression analysis and associations with insulin-stimulated LGU, as a measure of muscle insulin sensitivity. We observed that 221 proteins had different levels in insulin sensitive and insulin resistant subjects (*P_adj_* < 0.05, FC > 1.25), and 93 proteins correlated with LGU (*P_adj_* < 0.05) (**Figure 2C, Table S5,6**). Of these proteins, 54 were both differentially regulated and associated with insulin sensitivity, including metabolic enzymes (ACOX3, ADO, CKB, GALK1, UROS), proteins that regulate phosphorylation (PAK1, PAK6, PPP3CB, DUSP29), ubiquitination (TNFAIP3, HERC6, LRRC41, FBXO7), methylation (METTL3), actin (DMTN, LIMCH1), transcription (SMAD2, VPS39, SMARCD3), and a subset of proteins highly expressed in red blood cells (HBG1, SPTA1, ANK1, EPB41, GLUT1). Levels of core insulin action proteins such as INSR and GLUT4 did not differ between IS and IR individuals and were not correlated with LGU (**Figure 2D-G**). Of the canonical insulin signalling pathway proteins obtained from the union of KEGG, GO and Reactome databases (**Table S7**), only 8/156 proteins had differing levels between IR and IS subjects (Padj < 0.05, FC > 1.25) (**Figure 2H**). Of these proteins, three were also associated with insulin-stimulated LGU (*P_adj_* < 0.05) (PAK1, SORBS1, SOS1). The remaining 90 proteins associated with LGU that are not currently part of the insulin signalling network provide opportunities to further delineate their roles (**Figure 2I**).

**Figure 2:**
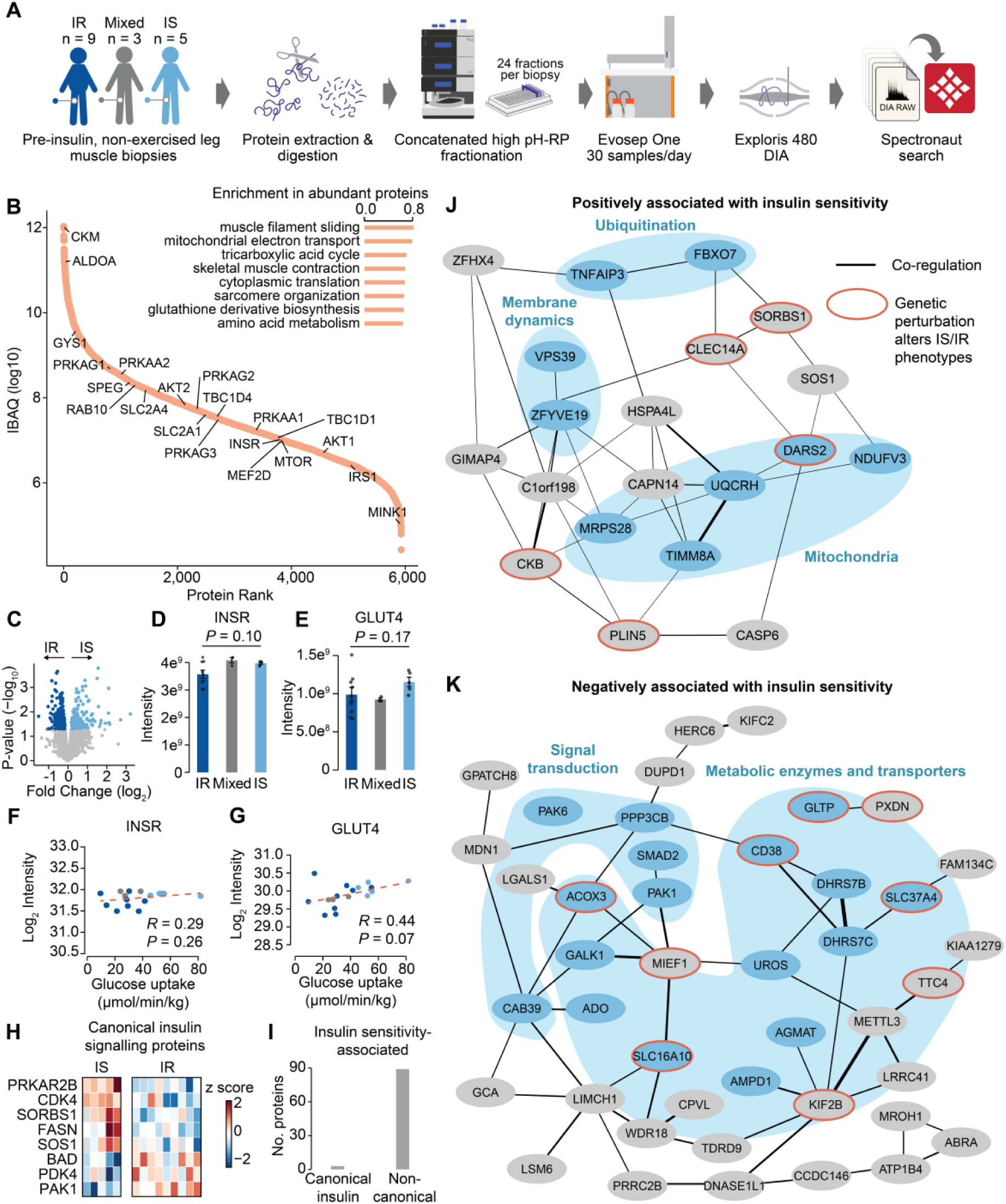
Insulin resistance involves altered levels of metabolic and signalling proteins, but not core insulin signalling proteins. **(A)** Schematic of proteome sample preparation workflow. **(B)** Ranked abundances of proteins in skeletal muscle. The top 8 most enriched non-redundant GO biological processes in muscle tissue proteins ranked by abundance (*P_adj_* < 0.05, two-sided Kolmogorov–Smirnov test). **(C)** Volcano plot of protein levels in insulin resistant and insulin sensitive skeletal muscle biopsies. Abundance of **(D)** INSR and **(E)** GLUT4 in skeletal muscle biopsies. Levels of **(F)** INSR and **(G)** GLUT4 correlations with insulin-stimulated LGU. **(H)** Heatmap of canonical insulin signalling proteins compiled from KEGG, GO and reactome insulin pathways that were differentially regulated between IR and IS subjects (*P_adj_* < 0.05 and FC > 1.25). **(I)** Number of proteins associated with steady state leg glucose uptake during a hyperinsulinemic euglycemic clamp (*P_adj_* < 0.05), from canonical insulin signalling and non-canonical proteins. Co-regulation networks of proteins (**J**) positively and (**K**) negatively associated with steady state leg glucose uptake during an insulin clamp. The width of the edges indicates the degree of co-regulation. Proteins with an orange outline have either a mouse knockout that causes a glucose or insulin-related phenotype^80^, a CRISPR KO in HeLa cells that alters insulin stimulated GLUT4 translocation^81^ or a SNP associated with type 2 diabetes in humans^50,62^. A dense module of blood proteins in the positively correlated group are not shown for clarity. *P*-value adjustments were calculated from permutations 1,000 times.

To contextualise the proteins associated with insulin sensitivity, we built co-regulation networks (**Figure 2J, K**). Proteins with common cellular localisations or functions clustered together, including a subset of mitochondrial proteins within the positively correlated group, which is consistent with previous reports^32^. This subset of mitochondrial proteins is involved in mitochondrial protein synthesis (MRPS28, DARS2), chaperones (TIMM8A), assembly factors (NDUFAF3) and complex III of electron transport (UQCRH). However, a small number of mitochondrial proteins were also negatively associated with LGU, including MIEF1, which regulates mitochondrial fission. We observed a dense cluster of red blood cell-specific proteins in the positively correlated network, which suggests that despite no changes in bulk blood flow between subjects (**Figure S1A**), microvascular flow may be higher in insulin sensitive individuals, which has previously been proposed^33^. Positively associated proteins with LGU also included proteins involved in ubiquitination (FBXO7, TNFAIP3), and membrane dynamics (VPS39, ZFYE19), which may influence GLUT4 trafficking in response to insulin. The proteins that were negatively correlated with insulin sensitivity included metabolic enzymes (GALK1, AMPD1, AGMAT, ADO, ACOX3, UROS), transporters (GLTP, SLC37A4, SLC16A10) and signalling components (SMAD2, PAK1, PAK6, CAB39, PPP3CB) (**Figure 2K**). To provide information about potential directionality within the co-regulation network, we overlaid published evidence from mouse knockouts, CRISPR screens or human genetic associations^34–37^. For example, PLIN5 knockout mice had insulin resistant skeletal muscle and increased ceramide levels, while muscle specific PLIN5 overexpression was metabolically protective^38,39^. We observed that PLIN5 levels were positively correlated with insulin sensitivity, so PLIN5 may similarly impact insulin sensitivity in human skeletal muscle, potentially via regulating triacylglycerol lipolysis and ceramide levels.

### Insulin resistance involves a selective defect in a subset of insulin-regulated signalling

Since insulin-responsive leg glucose uptake is a dynamic process, the difference between IS and IR may be driven by changes in intracellular signalling in muscle. To delineate such differences, we measured global phosphoproteomes of 114 skeletal muscle biopsies (6 biopsies per subject, n=19) (**Figure 3A**). Employing recent advances in MS technology we used data-independent acquisition (DIA) to quantify >26,000 phosphopeptides and >13,000 phosphopeptides across at least half of the samples measured (**Figure 3B, Figure S3A, Table S8**). Phosphoproteomes were highly reproducible with a mean correlation-coefficient of 0.91 (**Figure S3B**). As we have previously observed, individual subjects had their own phosphoproteome “fingerprints”, with phosphoproteomes clustering most strongly by individual (**Figure S3C**)^19^.

**Figure 3:**
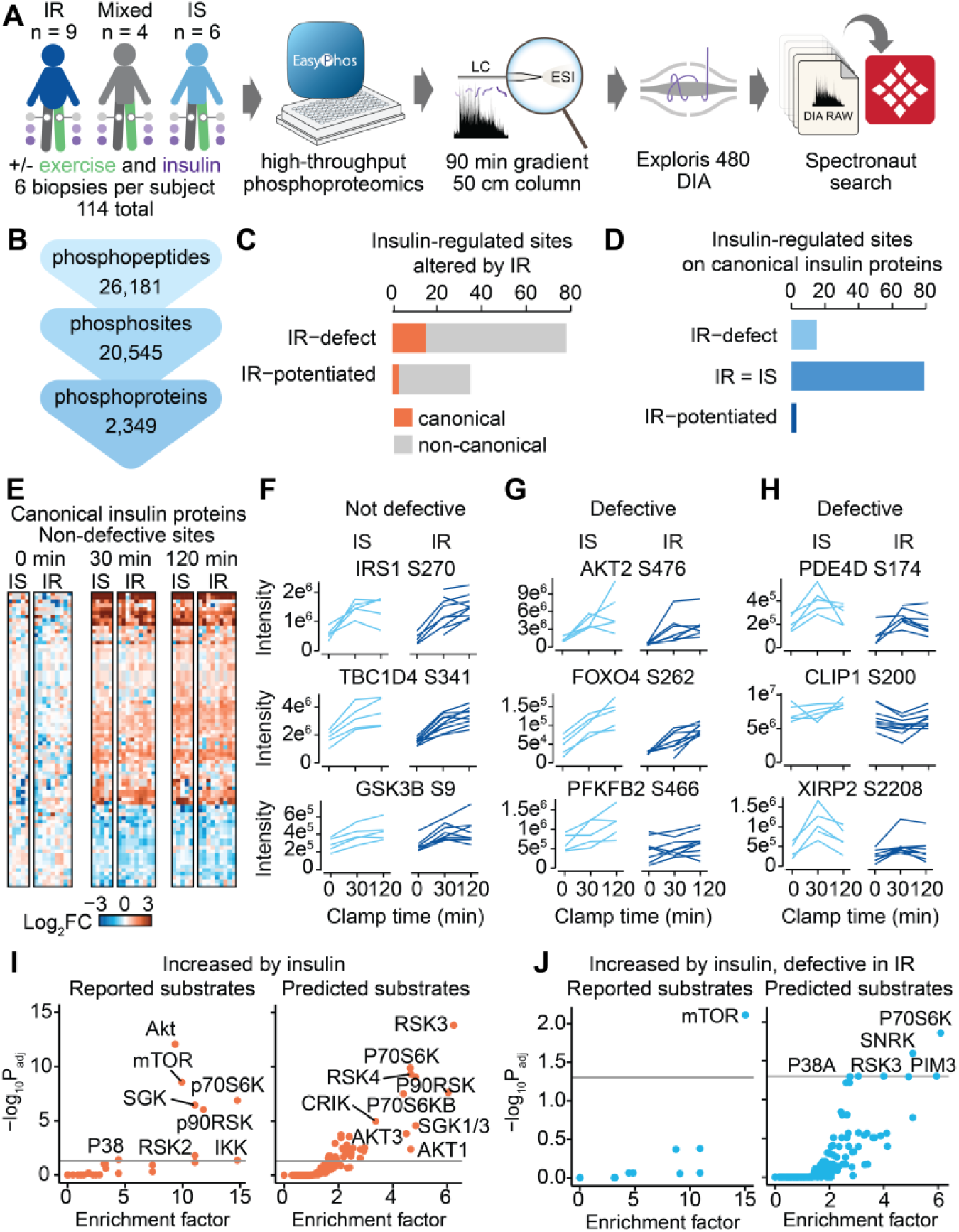
The phosphoproteome response to insulin is selectively defective in insulin resistant subjects. **(A)** Experimental workflow for global phosphoproteomics of skeletal muscle biopsies. **(B)** Quantification of the phosphoproteome. **(C)** The number of phosphosites that are regulated by insulin and either defective or potentiated in IR individuals coloured by whether that phosphosite is on a protein that is classified as canonical or non-canonical in insulin signalling. **(D)** The number of insulin-regulated phosphosites on canonical insulin signalling proteins separated into sites that are equivalent, defective, or potentiated in IR compared to IS subjects (n = 9 vs n = 6). **(E)** Phosphosites on canonical insulin signalling proteins that have not defective, including profiles of specific examples in **(F).** Example profiles of phosphosites with defective regulation by insulin in IR subjects, occurring on **(G)** canonical insulin signalling proteins and **(H)** proteins external to the canonical insulin signalling network. Enrichment of reported kinase substrates and predicted substrates by kinase library motifs in **(I)** all phosphosites increased by insulin and **(J)** phosphosites increased by insulin and defective in insulin resistant individuals (one-sided Fisher’s exact test, BH P-value adjustment).

We identified 1,794 phosphosites regulated by insulin, IR vs IS, or the interaction between these factors in biopsies from the non-exercised legs (*P*_adj_ < 0.05, FC > 1.25) (n = 9 IR vs n = 6 IS) (**Table S9**). Intriguingly, most of the sites that were significantly different between IR and IS subjects (749 sites) were not regulated by insulin (77 regulated in both comparisons, see “Defective phosphorylation in insulin resistance” below). Our observation that most differences between IR and IS are independent of the response to insulin is consistent with previous observations in cell culture^13^. Rewiring of the ground state proteome and phosphoproteome in IR muscle could be a feature of muscle IR that acts either in concert with or independently of the insulin-responsive sites.

To determine how insulin responses rather than the ground-state effects differ between IR and IS subjects, we defined “defective” signalling as phosphosites that are either regulated by insulin to a greater extent in IS than IR subjects, regulated in IR vs IS comparison, or have a significant interaction effect between insulin sensitivity and insulin (*P*_adj_ < 0.05, FC > 1.25). Notably, we also observed IR-“potentiated” signalling, whereby the effect of insulin was greater in IR compared to IS individuals. Overall, we found that more phosphosites had a defective rather than potentiated insulin response in IR individuals (78 vs 35) (**Figure 3C**).

#### Canonical insulin signalling

Insulin-regulated sites were enriched with sites found on canonical insulin signalling proteins (*P* < 2.2e^−16^) (**Figure S3D**). For example, we observed increases in the phosphorylation of AKT2 S474 (30 min median 6.8-fold, 120 min median 9.0-fold) and the doubly phosphorylated peptide containing S6 S236 (30 min median 26.0-fold, 120 min median 21.5-fold). Notably, the sites that differed between IR and IS subjects were not enriched for canonical insulin signalling (*P =* 0.39) and known canonical insulin signalling sites did not differ between IR and IS subjects (**Figure 3D-F, Figure S3D-F**). Although 19% of the IR defective sites mapped to proteins known to be involved in insulin signalling, most of these were phosphosites not implicated in canonical insulin signalling. For example, AKT2 S476 was defective in IR individuals (**Figure 3G**), but the canonical insulin signalling phosphosites on these proteins including AKT2 S474 were not defective (**Figure S3F**). The remaining 81% of phosphosites with a defective insulin response in IR subjects were not found on canonical insulin signalling proteins (**Figure 3C**). These include proteins with roles adjacent to insulin action such as PDE4D, which regulates cyclic AMP levels, XIRP2, which regulates actin polymerisation, and microtubule transport via CLIP1 (**Figure 3H**). Proteins with uncharacterised functions including C4orf54 also had defective phosphosites, representing opportunities to further expand our knowledge of the insulin signalling network and how it goes awry in insulin resistance.

#### Defective phosphorylation in insulin resistance

Of all insulin-regulated sites (54% sites up-regulated, 46% down-regulated), more sites that were defective in IR were increased by insulin than decreased in IS subjects (85% sites up-regulated, 15% down-regulated) (**Figures S3G, H**). Permutation testing confirmed that this difference was not driven by insulin up-regulating slightly more sites (*P* = 0.001) (**Figure S3I**). Based on these data, we conclude that IR in muscle appears to principally involve a defect in insulin-regulated phosphorylation, and not insulin-regulated dephosphorylation.

We next examined the known and predicted kinases that regulate insulin-mediated increases in phosphorylation. The insulin-regulated phosphoproteome was enriched in kinases involved in canonical insulin signalling including AKT, mTOR, p70S6K, SGK, p90RSK (**Figure 3I**). Most notably, the defective insulin-responsive sites in IR were enriched for mTOR substrates (*P*_adj_ = 0.007) (**Figure 3J**). These included PATL1 S179, S184, MAF1 S75, AMPKα2 S377 and MKNK2 S74, which are all rapamycin-sensitive (**Figure S3J**). Two reported AKT substrates and key regulatory sites were classified as defective (FOXO4 S262 and PFKFB2 S466) (**Figure 3G**). However, this did not reflect a general defect in AKT activity since 17 AKT substrates were increased by insulin and were equivalent between IR and IS individuals. This confirms previous reports that a uniform decrease in AKT activity does not occur during IR^40^. Alterations in the activity of AKT towards specific substrates, defective alternate kinases, or alterations to the activities of the phosphatases acting on the two reported AKT substrates with defective insulin responses may explain this observation.

Limited annotation of the substrates of kinases may obscure less well-studied kinases from being implicated in insulin resistance, since only 11/76 defective sites have a reported upstream kinase (**Figure S3K**). Testing for kinase enrichment based on predictions using substrate motifs expanded insulin-stimulated sites to 38, with 5 of these enriched in IR defective sites (**Figure 3I, J**). Sucrose non-fermenting AMPK-related kinase (SNRK/SNARK) has 0 reported substrates in PhosphoSitePlus and only one published substrate (PPP2R5D S80)^41^. However, based on motif predictions, SNRK activity may be defective in IR (**Figure 3J**). SNRK is an AMPK kinase family member and is activated by LKB1. In mouse skeletal muscle, overexpression of a dominant negative SNRK mutant reduced contraction- but not insulin-stimulated glucose transport in response to a glucose bolus^42^. SNRK forms a complex with CAB39, which we observed to be negatively correlated with insulin sensitivity at the protein level (**Figure 2K**)^43^. Investigating the role of SNRK in skeletal muscle in insulin resistant contexts may further clarify its role in metabolism. Consistent with the enrichment of reported mTORC1 substrates among sites defective in IR, p70S6K, which is directly downstream of mTORC1, was the most enriched predicted kinase in defective sites (**Figure 3J**). When examining reported mTORC1 substrates, we noticed an outlier individual with consistently lower phosphorylation of mTORC1 and p70S6K substrates (**Figure S3J**). This individual also had the second lowest GIR of all subjects (**Figure S3L**).

### Associating across the spectrum of muscle insulin sensitivity

The diversity of insulin sensitivity and phosphoproteome responses motivated us to utilize our recently described personalized phosphoproteomics approach to pinpoint signalling most likely to be important for insulin-stimulated glucose uptake in IR *vs* IS subjects^44^. Personalized phosphoproteomics utilises individual phenotypic and molecular diversity, rather than relying on discrete groupings (e.g. IR *vs* IS, or exercised *vs* rested) to link signalling with functional phenotypes. This approach provides a prioritised set of phosphosites that may be implicated in the biological function measured. Insulin-stimulated LGU greater in IS than IR subjects (**Figure 4A**). However, this phenotype was not discrete, but had high biological variance. Subjects classified as IS by HOMA-IR and GIR had a 1.8-fold range of LGU (**Figure 4B**). Incorporating all lean subjects with a HOMA-IR classified as IS, including the “IS” and “mixed” groups, encompassed a range of 6.4-fold. This is slightly higher than that observed even in IR participants, which had a 5.8-fold range of LGU. This diversity is obscured in discrete comparisons of both glucose uptake and phosphoproteomics measures, as opposed to analysis of continuous measures.

**Figure 4:**
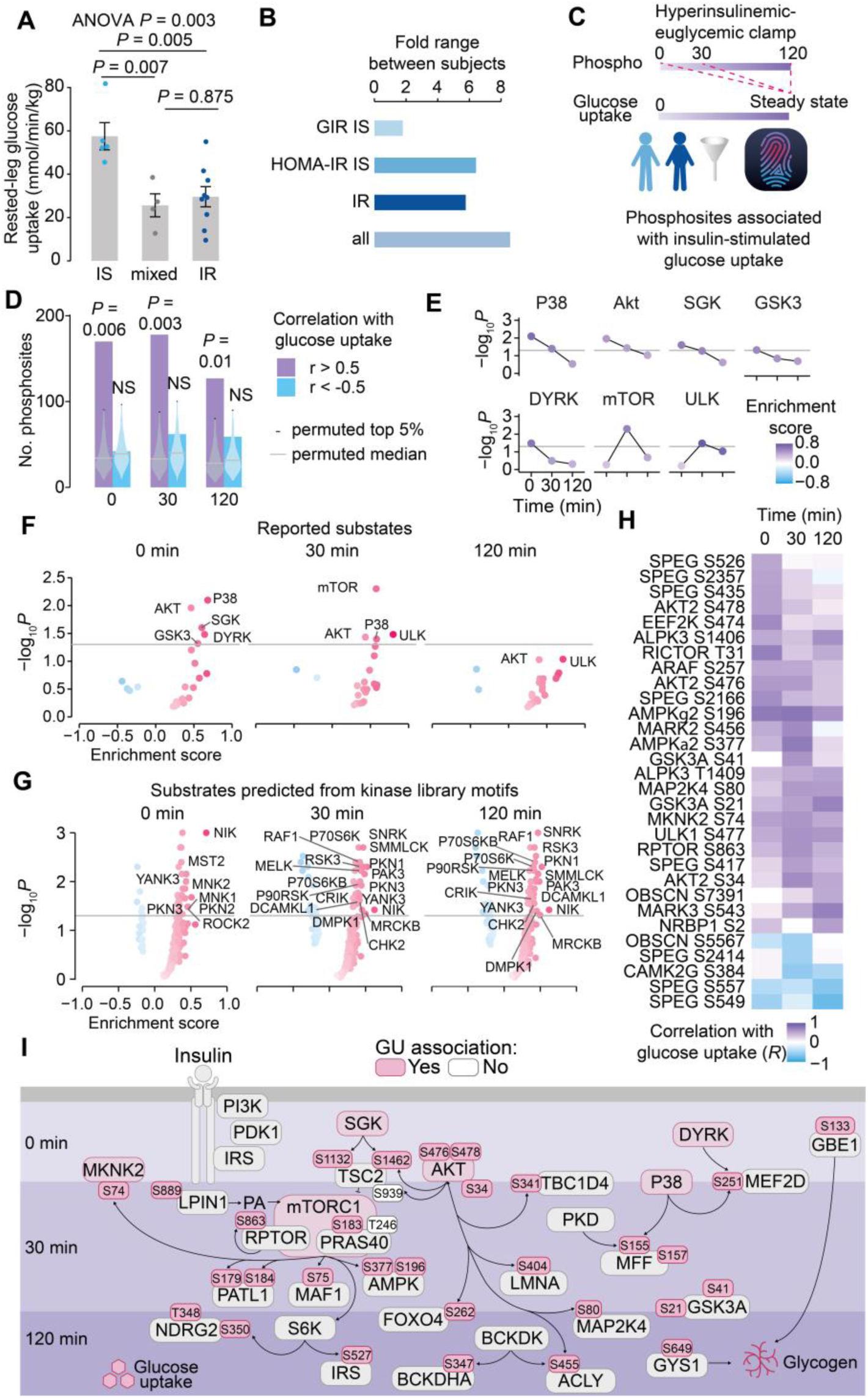
Personalized phosphoproteomics identifies signalling associated with defective glucose uptake in insulin resistant subjects. **(A)** Insulin-stimulated LGU in the non-exercised leg (mean of the 90, 105, 120 min insulin time points, ANOVA with Tukey HSD). **(B)** The fold ranges of LGU in the subject groups. **(C)** Schematic illustrating our temporally-shifted personalized phosphoproteomics approach, associating the phosphoproteomes of each time point with LGU. **(D)** The number of phosphopeptides that correlate with LGU with an |r| > 0.5 per timepoint. Permutations were performed 1,000 times for each timepoint, by shuffling the glucose uptake sample labels and associating these with all sites in the correlation analysis (significantly regulated in the rested leg ANOVA and fold range between max-min at that time point >= 1.5-fold). Empirical *P*-values were calculated from the permutations. Profiles of **(E)** significant and **(F)** all kinase enrichments based on reported substrates and **(G)** predicted substrates based off kinase library motifs, ordered by their correlation-coefficients (modified two-sided Kolmogorov-Smirnov test). **(H)** The associations of phosphosites on kinases with steady state glucose uptake (P_adj_ < 0.05 in at least one timepoint). **(I)** Signalling diagram of selected sites of interest with phosphosites and kinases coloured if associated with glucose uptake at the timepoint represented in the layers (*P_adj_* < 0.05). P-values were adjusted by calculations from permutations 1,000 times.

Here, we extended our personalized phosphoproteomics approach by including a time-shifted component to prioritise signalling potentially occurring prior to and upstream of measured phenotypes. To this end, we associated non-exercised leg phosphosite profiles from each timepoint of the insulin clamp (0, 30, 120 min) with steady-state glucose uptake in the non-exercised leg, referred to as LGU (**Figure 4C**). Association analyses were performed on phosphosites if they were significantly regulated and differed between individuals by at least 1.5-fold at that timepoint, to select for phosphosites with sufficient inter-subject heterogeneity. This analysis revealed 512 phosphosites associated with LGU (*P*_adj_ < 0.05) in at least one time point (**Figure 4D, Table S10**). Permuting the subject LGU 1,000 times and testing their associations with signalling revealed that positively-associated phosphosites (*R* > 0.5) were greater than expected by chance. In contrast, negatively-associated phosphosites (*R* < -0.5) were equivalent to what would be expected by chance. This is consistent with our observation that most of the phosphosites with defective insulin responses in IR subjects are increased in abundance by insulin in IS subjects (**Figure S3G**). We were able to expand the identification of sites potentially involved in insulin-stimulated LGU, as we identified 480 sites of interest in this analysis, compared with discrete comparisons (overlap of 32, **Figure S4A**).

Introducing temporal resolution to an insulin-stimulated LGU association may differentiate biological stages and distinguish signalling that is less likely to occur due to reverse causation (although confounding remains a possibility). The 0-minute timepoint may reflect ground-state differences in the muscle proteome or signalling architecture that could influence an individual’s ability to respond to insulin. The 30-min timepoint could reflect insulin-responsive signalling that promotes glucose uptake, and 120-min may involve signalling that either maintains glucose uptake or feedback regulation responding to increased glucose flux into muscle cells.

Representing the ground-state architecture prior to the clamp (0 min), phosphosites positively associated with LGU measured at the end of the clamp were enriched with reported substrates of p38, AKT, SGK, DYRK and GSK3, and many predicted substrates of kinases (**Figure 4E-G**). We observed the pathway “protein localisation to the PM” was enriched in proteins with positively associated sites at the 0-minute timepoint, including TBC1D4 S341 and CLASP2 S529 (**Figure S4B**). CLASP2 knockdowns reduce GLUT4 localisation at the plasma membrane^45^. Although broader knowledge of kinase-substrate relationships is required to comprehensively label upstream kinases, it is notable that IS individuals have higher levels of phosphosites observed on proteins involved in GLUT4 translocation in their ground-state. This may prime the glucose uptake machinery in IS subjects.

We observed a decline in the correlation of reported AKT substrates with LGU over the time-course of the clamp, including GLUT4 regulators such as TBC1D4 S341 (0 min *R* = 0.57, *P_adj_* = 0.013, 30 min *R* = 0.53, *P_adj_* = 0.034, 120 min *R* = 0.21, *NS*) (**Figure 4F**). However, a small subset of AKT substrates comprising of six sites remained associated with LGU even at 120 min, including GSK3α S21 (120 min *R* = 0.70, *P_adj_* = 0.003) and FOXO4 S262 (120 min *R* = 0.52, *P_adj_* = 0.033) (**Figure S4C**).

Phosphorylation of AKT2 on S34, a site without a known function, was increased with insulin. AKT2 S34 levels at the 30 and 120 min timepoints positively correlated with LGU (**Figure S4D**). A previous report demonstrated that ceramide causes PKC-mediated phosphorylation of AKT3 S34 to inhibit AKT3, with the readout of AKT activity being S473 phosphorylation^46^. In contrast to this, we observed higher AKT2 S34 phosphorylation in more insulin sensitive subjects, which would be inconsistent with ceramide-induced activation. Given that AKT substrates are strongly phosphorylated in response to insulin, we could find no evidence that S34 phosphorylation inhibits AKT activity,^46^at least in this context. S34 occurs in the plekstrin homology domain of AKT that is required for PIP_3_ binding at the plasma membrane. The small subset of glucose uptake associated AKT substrates at 30 and 120 min may be localized to a specific subcellular compartment if S34 promotes the release of AKT from the plasma membrane.

The insulin-responsive signalling observed at 30 min during the clamp contributed the greatest number of phosphosites associated with muscle glucose uptake, compared to 0 or 120 min (**Figure 4D**). Phosphorylation of mTOR substrates at 30 min were the most strongly associated with LGU of any substrates detected (*P* = 0.006). Some of the key mTOR substrates that were detected here include PATL1 S184, MAF1 S75 and PRKAA2 S377. Consistent with mTORC1 substrate enrichment, regulatory phosphosites on mTORC1 measured at 30 min were associated with LGU, including the mTORC1 activating site RPTOR S863 (*P*_adj_ = 0.011, *R* = 0.616), and the mTORC1-catalysed phosphorylation of PRAS40 (AKT1S1) S183 that promotes mTORC1 substrate phosphorylation (*P*_adj_ = 0.013, *R* = 0.618). The AKT site on PRAS40 (T246) was not associated with LGU at any time point (0 min *R* = -0.25, 30 min *R* = 0.05, 120 min *R* = -0.23, *P*_adj_ > 0.05). The SGK substrate TSC2 S1132 and dual SGK/AKT substrate T1462 inhibit TSC2 to activate mTORC1^47,48^. Ground-state levels of these phosphosites were positively associated with LGU, so SGK and/or AKT may prime mTORC1 to respond more strongly to insulin in subjects with greater insulin sensitivity at 30 min.

We observed that phosphorylation of the mTORC1 substrate AMPKα2 S377 was positively associated with LGU (**Figure S4E**). We previously identified this site during the exercise-mediated potentiation of insulin sensitivity in lean, IS individuals^44^. Signalling at this site also varies across the spectrum of insulin resistance. In addition to S377 on the α2 subunit of AMPK, we identified a phosphosite on the γ2 phosphosite of AMPK, S196, with 30 min levels that also correlate with LGU (**Figure 4H-I, S4F**). Concomitant with the regulated AMPK phosphosites, we observed the enrichment of proteins involved in the “response to glucose starvation” at 30 min into the clamp (**Figure S4G**). In cultured adipocytes, energy charge drops in the first 20 min of insulin stimulation as the signalling activates anabolism prior to glucose entering the cell^49^. If this also occurs in skeletal muscle during a clamp, this “pull” of glucose into the cell during early clamp timepoints may involve AMPK regulation.

Despite being at a steady state of glucose uptake, the muscle biopsy sampled at the 120 min timepoint had far less insulin responsive phosphosites associated with LGU than those sampled at 0- or 30-minutes (**Figure 4D**). Notably, there was no enrichment of kinases with known substrates observed at this timepoint. This may reflect poor kinase annotation^24^, particularly with respect to late-stage insulin signalling, as phosphorylation sites on kinases with few reported substrates including MARK3 and ALPK3 were positively associated with LGU at this time. It is possible that the yet unknown substrates of these kinases and other kinases may also track with glucose uptake. An intron variant in *MARK3* is associated with type 2 diabetes^50^. ALPK3 also has a potential role in glucose uptake, as knockdown of ALPK3 (MIDORI) in HeLa cells increased basal and IGF1-stimulated GLUT4 translocation to the plasma membrane^51^. Expanding the repertoire of known kinase-substrate relationships will therefore further clarify the regulatory mechanisms of insulin-stimulated glucose uptake.

### Prior exercise primes a subset of the defective phosphorylation in insulin resistance

Prior exercise potentiated insulin-stimulated glucose uptake (*P* = 5.2e^−7^), and regulated 443 phosphosites (BH *P_adj_* < 0.05) (**Figure 5A, Table S11**). As in the non-exercised leg, the magnitude of insulin sensitization by exercise varied substantially across individuals. The greatest magnitude of difference in insulin-stimulated LGU between the exercised and non-exercised legs was 2.63-fold and the lowest was 0.98-fold (no response). We associated phosphoproteomes from each time point with the insulin-stimulated LGU as above. Here, to identify signalling associated with the potentiation of insulin-stimulated LGU by prior exercise, we first performed repeated measures correlations to associate the difference in phosphorylation between the non-exercised and exercised legs of each subject with the change in insulin-stimulated LGU between legs (**Figure 5B, Table S12**)^52^. As for the rested leg associations, we observed a similar shift towards positive correlations, however, the 120 min timepoint had equivalent positively and negatively associated phosphosites (**Figure 5C**). Inhibitory site S641 on GYS1 was the most strongly correlated site with a reported function at this time point (**Figure S5A**). Their negative association with LGU fits the positive link between insulin stimulation and exercise-potentiated glycogen synthesis^53^.

**Figure 5:**
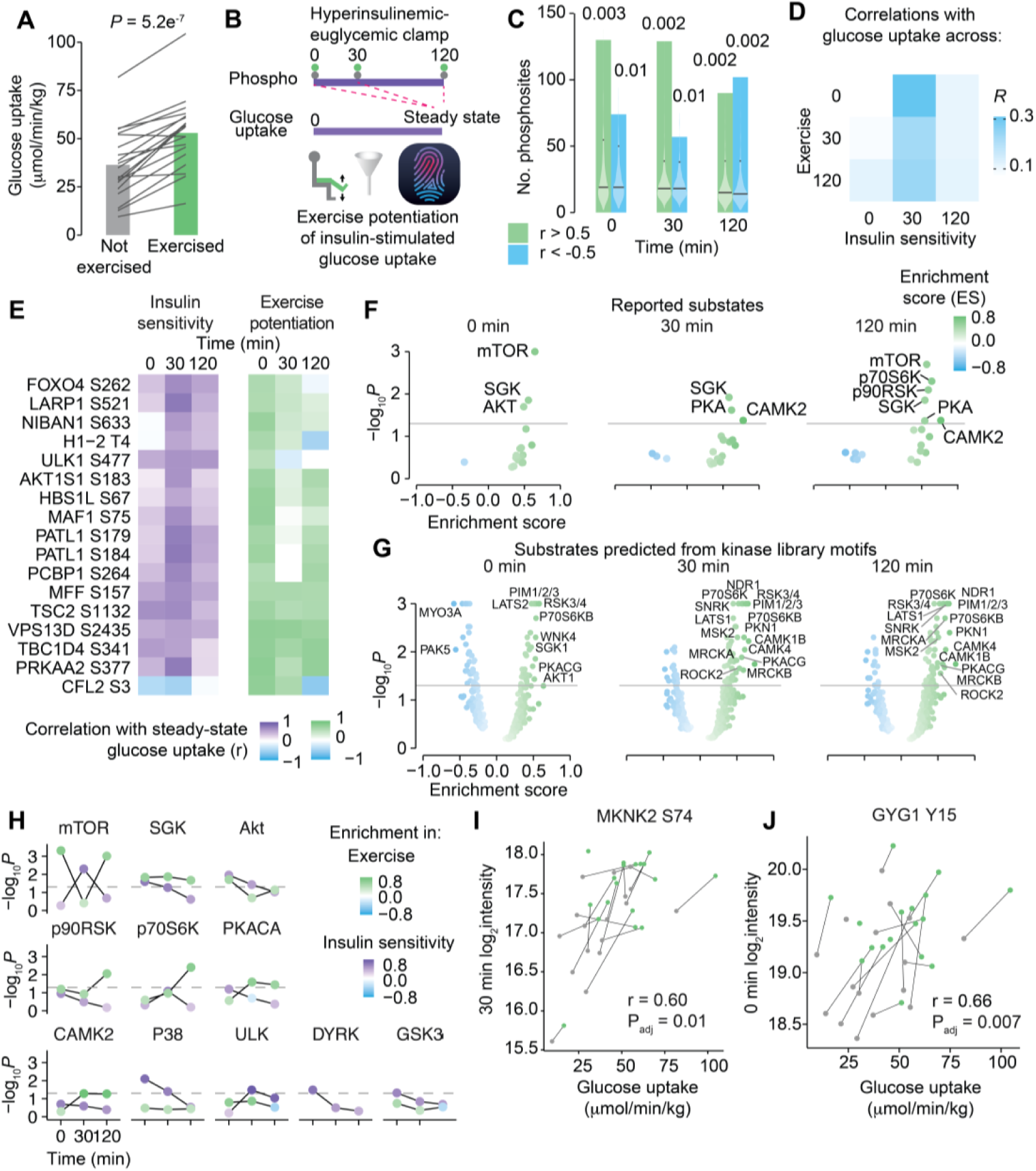
Exercise primes phosphosites that are associated with steady state glucose uptake in insulin sensitivity. **(A)** Steady-state leg glucose uptake into the non-exercised and exercised legs of each subject (paired t-test). **(B)** Schematic demonstrating repeated measures correlations to associate the phosphoproteomes of the non-exercised and exercised legs with steady state leg glucose uptake. **(C)** The number of phosphopeptides that correlated with steady-state glucose uptake with an |r| > 0.5 per timepoint. Permutations were performed as in **4D**. **(D)** Comparison of the associations with the degree of insulin sensitivity performed in Figure 4D labelled “Insulin sensitivity” with the associations with the potentiation of glucose uptake by exercise, labelled “Exercise”. **(E)** Heatmap of the phosphosites that were associated with steady state glucose uptake across insulin sensitivity at 30 min and exercise potentiation at 0 min (*P*_adj_ < 0.05). P-values were adjusted by permutation 1,000 times. Kinase enrichment on **(F)** reported substrates and **(G)** predicted substrates based on kinase library motifs, ordered by their correlation-coefficients with steady state glucose uptake. **(H)** Profiles of any significantly enriched kinase based on reported substrates when associating across insulin sensitivity or exercise potentiation, comparing their enrichments (*P_adj_* < 0.05). *P*-value adjustments were calculated from permutations 1,000 times. **(I)** MKNK2 S74 and **(J)** GYG1 Y15 phosphorylation and steady state glucose uptake. Grey represents the non-exercised leg and green represents the exercised leg, with lines connecting samples from the same subject.

We next compared the phosphorylation sites that were associated with exercise-potentiated insulin responses versus associations of ground-state insulin sensitivity across subjects. Of all time points, phosphosites at the 0-minute timepoint of the insulin clamp that were associated with sensitization by exercise were most similar with phosphosites at the 30-minute time point of the insulin clamp that were associated with rested leg insulin sensitivity (*R* = 0.31, *P*_adj_ = 7.9e^-11^) (**Figure 5D**). Exercise recovery may therefore alter the ground-state signalling to shift the cell state closer to insulin-stimulated cells in the more insulin sensitive subjects. This signalling involves an overlap of 17 sites including the mTORC1 substrates PATL1 S184, AMPKα2 S377, MAF1 S75, and PRAS40 S183 (**Figure 5E**). The SGK substrate TSC2 S1132 that activates mTORC1 and the AKT substrates FOXO4 S262 and TBC1D4 S341 were also associated with LGU. The shift of the common associated signalling from 30 in the rested leg to 0 minutes for exercise was also evident in kinase enrichment, as mTOR substrates were enriched in positively associated phosphosites at the 0 min time point in the exercise sensitization associations, equivalent to the 30 min time point of the insulin sensitivity associations (**Figure 5F**). AKT and SGK remained enriched at the 0 min time point, as in the insulin sensitivity associations (**Figure 5G, H**). Since glucose uptake was not higher in the ground state exercised leg, this potentiation of signalling was not sufficient to promote increased glucose uptake but may rather be involved in priming the potentiated response to insulin.

Comparing phosphosites associated with insulin sensitivity across subjects and those associated with insulin sensitization by exercise at any time point revealed 49 shared sites (**Figure S5B**). Mitochondrial fission was represented via VPS13D S2435 and MFF S157, the phosphorylation of which were positively correlated with steady state glucose uptake in both comparisons. MFF S157 is a reported regulatory site, and the highest motif score is for p70 S6K^54^. The S455 phosphosite on the metabolic enzyme ATP citrate lyase (ACLY) is a reported regulatory phosphosite targeted by AKT or BCKDK^55,56^. ACLY catalyses the formation of the lipid malonyl-CoA, an allosteric inhibitor of fatty acid oxidation. Reduced malonyl-CoA content in skeletal muscle during exercise recovery has previously been observed to correlate with improved insulin-stimulated glucose uptake in skeletal muscle^57^. This anabolic shift may be a consequence or cause of increased glucose uptake, or an independent arm of insulin signalling that is also more insulin sensitive. Kinases with few reported substrates were also represented, as the inhibitory phosphosite S74 on MKNK2 was positively associated with LGU in non-exercised legs, and in exercise-mediated insulin sensitization (**Figure 5I**). MKNK2 S74 is an mTORC1 substrate, and loss of this phosphorylation site increases cell size via reciprocal regulation onto mTORC1^58^. MKNK2 KO mice are more insulin sensitive and glucose tolerant on a high fat diet than WT mice^59^. The inhibitory phosphorylation of MKNK2 by mTORC1 may influence insulin sensitivity in humans. Delineating the substrates of MKNK2 may further clarify its role in metabolism.

Most of the signalling associated with the exercise-mediated potentiation LGU was divergent from the signalling associated with insulin sensitivity in the non-exercised legs. It is therefore possible that except for the shared subset of sites including mTORC1 substrates, insulin resistance may involve different signalling mechanisms to those promoted in the insulin sensitization by exercise. We identified 331 phosphosites correlated with LGU potentiation by exercise that were not associated with insulin-stimulated LGU in the non-exercised leg at any time point. These include S318, an additional regulatory phosphosite on TBC1D4 and the regulatory site S319 on PCYT1A, an enzyme that catalyses phosphatidylcholine biosynthesis (**Figure S5A**). We also observed that the regulatory sites on the kinases PRKAR1A S77, MAPK1 Y187 and RPS6KA3 S369 were positively associated with LGU potentiation by exercise (**Figure S5A, C**). PRKAR1A is the regulatory subunit of PKA and the magnitude of S77 phosphorylation at 0 and 30 min correlated positively with insulin sensitization by exercise. PKA substrates were enriched in positively correlated phosphosites at 30 min, which is surprising since insulin inhibits PKA. Of the substrates positively associated with insulin sensitization by exercise, 75% are also reported to be substrates of other kinases including AKT and SGK, so the upstream signal may be mixed. Novel phosphosites were also identified, including Y15 on glycogenin-1 (GYG1), which uniquely correlated with insulin sensitization by exercise, not insulin sensitivity in non-exercised legs (**Figure 5J**). GYG1 forms the core of glycogen and acts as a substrate for glycogen synthase by glucosylating itself. Y15 is immediately adjacent to an amino acid that is a missense variant causing polyglucosan myopathy (A16P), indicating that this may be a functionally important region of this protein^60^. At 0 min, phosphorylated GYG1 Y15 was higher in the exercised legs, and positively correlated with LGU. GYG Y15 phosphorylation may have a role in the synthesis of glucose into glycogen post-exercise.

### Phosphoproteomics contextualises disease-modifying genetic variants

Phosphoproteome associations prioritise phosphosites of potential interest but do not infer causation. Identifying phosphorylated proteins with a genetic variant associated with glycaemic traits would therefore increase confidence of the potential biological role of that phosphosite. Filtering phosphoproteomics findings for candidate genes from genome wide association studies of relevant phenotypes has successfully prioritised candidates previously^61^. We filtered our phosphoproteomes for proteins encoded by the genes nearest to published genetic variants associated with type 2 diabetes, fasting glucose, HbA1c, fasting insulin or 2hr glucose during an oral glucose tolerance test^50,62^. This analysis revealed 246 total sites on 58 proteins encoded by genes nearest T2D and glycaemic variants. Of these, 32 sites on 17 proteins were associated with LGU the continuum of insulin sensitivity or exercise-mediated insulin sensitization (*P*_adj_ < 0.05) (**Figure 6A**). These include proteins with known roles in insulin signalling (IRS1, IRS2), and metabolism (PFKM, AMPD3, ACSL1). We also identified proteins involved in cellular signalling that have not yet been characterised in the response to insulin (MARK3, ARHGAP1, MINDY1, HBS1L), representing candidate proteins potentially involved in insulin-stimulated glucose uptake.

**Figure 6:**
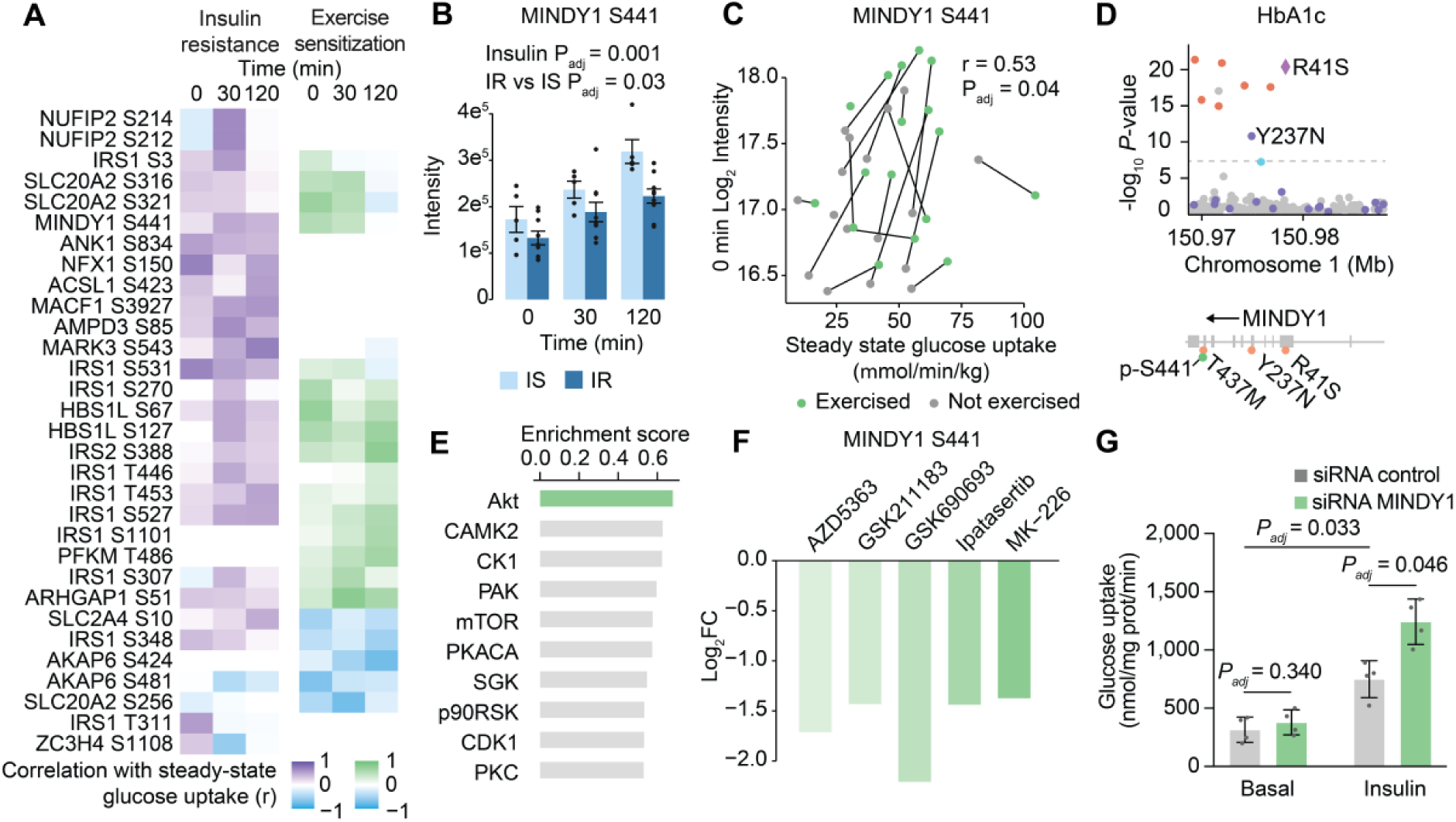
MINDY1 is downstream of Akt, is a negative-regulator of insulin-stimulated glucose uptake and contains a missense variant that is protective for type 2 diabetes in humans. **(A)** Phosphosites on proteins that are the nearest gene to a type 2 diabetes or glycaemic phenotype genetic variant, which are associated with steady state glucose uptake in at least one timepoint (*P*_adj_ < 0.05). *P*-value adjustments were calculated from permutations 1,000 times. **B)** Abundance of MINDY1 S441 in the non-exercised leg, comparing IS and IR groups. **(C)** Repeated measures correlation of MINDY1 S441 in the non-exercised and exercised leg at 0 min with the steady state leg glucose uptake. **(D)** Locus zoom plot of the region encoding *MINDY1*, indicating genetic variants and their significance of associations with HbA1c in a meta-analysis of genome wide association studies in the T2D Knowledge Portal. Chromosome coordinates are in GrCh37. **(E)** Enrichment of kinase substrates in all sites ordered by their co-regulation with MINDY1 S441. **(F)** MINDY1 S441 phosphorylation in BT-474 cells after treatment of with five different AKT inhibitors. This data was published in the supplementary material of^69^. **(G)** 2-[^3^H]deoxyglucose uptake into rat L6 myotubes treated with 100 nM insulin for 20 minutes with or without knockdown of *Mindy1*. Two-way ANOVA with Holm-Šídák’s multiple comparisons test correction. Error bars represent mean +/- SEM.

HBS1L S67 was regulated by insulin and differed between IR and IS individuals, the 30 min levels of HBS1L S67 were positively associated with LGU in non-exercised legs (*P*_adj_ = 0.048) and the 0 and 120 min levels correlated with insulin sensitization by exercise (**Figure S6A, B**). A variant nearest the HBS1L gene is associated with HbA1C^62^. HBS1L is a translation factor GTPase, and S67 and S127 occur in the domain of HBS1L that interacts with the ribosomal protein rpS3 at the mRNA entry site of the ribosome^63^. HBS1L S67 and S127 were co-regulated with p70 S6K and mTOR substrates (**Figure S6C, D**). Treating HEK and HeLa cells with insulin and two different mTORC1 inhibitors reduced the abundance of HBS1L S67, confirming that this phosphorylation event is downstream of mTORC1 (**Figure S6E**). The role of HBS1L in translation is consistent with the biological processes regulated by mTORC1, which may be implicated in skeletal muscle insulin resistance.

Phosphorylation of MINDY1 (FAM63A) S441 was linked to insulin sensitivity in both ground-state and exercise contexts. MINDY1 S441 was regulated by insulin, differed between IR and IS individuals, MINDY1 S441 levels prior to the clamp positively correlated with insulin sensitization by exercise (**Figure 6B, C**). Non-coding and missense variants in MINDY1 are protective for T2D and HbA1c levels^64–66^. Mapping the genetic coordinates to protein sequences using VarMap^67^, we identified the missense variants as R41S (GrCh37 chr 1 position 150,974,971 ref T alt A, HbA1c P = 4.69e^−21^ β = -0.015), Y237N (GrCh37 chr1 position 150,972,959 ref A alt T, HbA1c P = 1.91e^−11^, β = -0.049, T2D P = 5.8e^−8^ odds ratio = 0.82) and T437M (GrCh37 chr1 position 150,970,119 ref G alt A, T2D P = 6.16e^−6^ odds ratio = 0.54) (**Figure 6D**). The T437M variant is rarer than the other missense variants with a minor allele frequency of 0.0007 in gnomAD compared to 0.01 and 0.002 for R41S and Y237N, respectively^68^. The Y237N variant was previously included in a clustering analysis with other T2D-associated variants by metabolic traits, grouping Y237N into the “insulin action” group, rather than insulin secretion or dyslipidaemia^64^. Given the role of skeletal muscle in insulin-stimulated glucose disposal, this finding would be consistent with MINDY1 influencing type 2 diabetes risk via muscle glucose uptake rather than other processes like insulin secretion. MINDY1 removes Lys-48 conjugated ubiquitin from target proteins, so it could be involved in proteasomal degradation in skeletal muscle. Insulin is known to regulate proteasomal activity but the mechanism for this effect is not clear.

As MINDY1 S441 does not have a reported upstream kinase, we leveraged the biological variability of individuals across different signalling modules. We identified co-regulated phosphosites to predict an upstream kinase of this phosphosite. AKT substrates were most enriched in co-regulated sites, and AKT1 is within the top ten kinases based on motif score from the kinase library (**Figure 6E, S6F**)^54^. A published phosphoproteomic resource of the inhibition of AKT with five different compounds identified reduced MINDY1 S441 with all tested AKT inhibitors (**Figure 6F**)^69^. MINDY1 was not regulated by rapamycin in HEK cells, suggesting that this phosphosite is not downstream of mTORC1 and S6K, which shares a similar motif to AKT (**Figure S6G**). These results do not prove MINDY1 S441 is a direct AKT substrate but provides evidence that it is downstream of AKT in cells. To further explore a potential functional role of MINDY1 in insulin-stimulated glucose uptake, we knocked down *Mindy1* in insulin responsive rat L6 myotubes. A 70% knockdown of *Mindy1* did not alter ground-state glucose uptake but caused a 1.7-fold potentiation of insulin action on glucose uptake (*P_adj_* = 0.046) (**Figure 6G, Figure S6H**). This is consistent with the earlier observation that missense variants, which are predicted to cause deleterious effects on MINDY1 function, are protective for type 2 diabetes. Our analysis of MINDY1 places a type 2 diabetes candidate gene within the insulin signalling network in human skeletal muscle and identifies a functional role for this protein in muscle insulin action.

## Discussion

In this study, we measured parallel and time-resolved human phenotypes, proteomes and phosphoproteomes, to explore how insulin resistance reshapes skeletal muscle responses to insulin, and how prior exercise mediates insulin sensitization through signalling. We observed that individuals displayed a continuum rather than discrete differences in insulin sensitivity, and that discordances between HOMA-IR and GIR could suggest tissue-specific insulin resistance in some individuals. Insulin resistant and insulin sensitive skeletal muscle biopsies had equivalent levels of core insulin action proteins, while proteins involved in membrane dynamics, signalling and metabolism differed between the groups, and tracked with the continuous range of muscle insulin sensitivity. Similarly, we observed that of the signalling responses to insulin that were defective in IR individuals, 81% occurred on proteins outside the canonical insulin signalling network, and most of these did not have reported upstream kinases or downstream functions. Of the characterised phosphosites, mTORC1 substrates were most defective in insulin resistance. A subset of AKT substrates were defective, suggesting potential substrate-specificity of AKT dysregulation in insulin resistance. Prior exercise partially counteracted insulin resistance, sensitizing the insulin-stimulated LGU of the insulin resistant subjects to the non-exercised levels of insulin sensitive subjects. The signalling activated by prior exercise was largely separate to insulin-regulated signalling, except for a subset of signalling activated at 30 minutes by insulin in non-exercised legs, that was primed prior to any insulin infusion by exercise. This subset included mTORC1 substrates, and uncharacterised phosphosites including MINDY1 S441. We observed that MINDY1 S441 was downstream of AKT, and we identified that MINDY1 inhibits insulin-stimulated glucose uptake in rat myotubes, which is consistent with *MINDY1* missense variants being protective for type 2 diabetes in humans^37^.

To reduce the likelihood of phosphosites being prioritised due to reverse causation, we extended our previously described personalized phosphoproteomics framework. Incorporating temporally shifted analyses identified signalling that was regulated prior to changes in glucose uptake during the insulin clamp. Although mTORC1 can be activated by glucose^70^, we observed that mTORC1 substrates measured at an earlier time point were correlated with steady state glucose uptake, indicating that reverse causation is less likely in this case. Our observation of priming of mTORC1 regulatory sites by AKT and SGK suggest that initial minor differences in these or other kinases could be propagated through mTORC1. The positive association of mTORC1 activity with insulin-stimulated glucose uptake is surprising, given reported models of mTORC1 promoting insulin resistance by activating p70 S6K, which phosphorylates IRS on inhibitory sites in mice^71^. We did not observe IRS inhibitory phosphorylation being lost in response to exercise or increased in insulin resistant subjects, so this does not appear to be a major mechanism of insulin in human skeletal muscle in these contexts. There is potential for mTORC1 to have tissue specific roles with conflicting effects on insulin resistance. The mTORC1 inhibitor rapamycin causes hyperglycaemia in humans primarily by impairing beta-cell function, and also by altering hepatic gluconeogenesis^72–74^. mTORC1 mediated the beneficial effects of FGF21 in adipocytes, including glucose uptake^75^. Similarly, a muscle-specific knockout of TSC1 resulting in constitutive mTORC1 activation caused mice to be more insulin sensitive than their wildtype littermates when fed a high fat diet, measured by an insulin tolerance test^76^. Due to the apparent tissue and context-specific outcomes of mTORC1 activity, inhibiting mTORC1 in an experimental paradigm similar to this study will be required to determine the role of mTORC1 in skeletal muscle insulin sensitivity.

To prioritize the phosphosites outlined in this resource, we identified phosphosites correlated with the phenotype of interest, and integrated alternate evidence about potential causal regulators. By integrating candidates from genetic studies of relevant phenotypic outcomes, we also contextualised disease-associated genes HBS1L and MINDY1 in the insulin signalling network. This is useful because genetic studies of insulin resistance and type 2 diabetes have identified hundreds of genetic variants associated with glycaemic traits^36,37^. Yet major challenges remain in pinpointing affected genes, the tissues they act in and their downstream cellular effects^77^. Since we observed signalling changes on these candidates in human skeletal muscle in a way that differed in insulin resistance, it is possible that these candidates mediate some of their disease risk through skeletal muscle. Since MINDY1 and HBS1L are also expressed in other tissues involved in metabolism including brain, adipose tissue and pancreas, measuring signalling in these tissues would clarify if these proteins are responsive to insulin or correlated with insulin sensitivity phenotypes in other contexts than skeletal muscle^78^. Given that missense variants with predicted loss of function of MINDY1 are protective for type 2 diabetes, and we observed that knockdown of *Mindy1* increases insulin sensitivity, it is possible that MINDY1 S441 phosphorylation downstream of AKT acts in an inhibitory manner to promote insulin-stimulated glucose uptake in a similar manner to the knockdown. S441 occurs near the CAAX box domain, which regulates membrane organisation, so phosphorylation of S441 could be regulating cellular localisation to influence MINDY1 function^79^. Future cellular studies investigating how phosphorylation and missense variants alter MINDY1 function will further clarify its role in insulin action.

### Study Limitations

The subjects in the current study were all males of European ancestry within a narrow age range. Future studies including women and a range of ancestries and ages will therefore broaden the generalisability of our findings. Future work would also benefit from engaging larger cohorts to increase power to detect differences. We utilised P-value adjustment by permutation in the current study to capture potential changes and observe trends for example kinase enrichment, however, our findings may also include false positives as this method is less stringent than other P-value adjustment methods (e.g. Bonferroni or Benjamini Hochberg). We have provided all our results as raw data, which enables comparative and meta-analyses when additional studies are published. It is possible that overweight status, rather than insulin resistance *per se*, could be responsible for some differences between subject groups prior to insulin stimulation. We aimed to focus in on this by associating the ground state phosphoproteomes with insulin sensitivity in each individual. In the future, testing an overweight insulin sensitive group may help further delineate these differences. We selected rat L6 myotube cells for our functional work on the role of MINDY1 in insulin-stimulated glucose due to their capacity for insulin responsive glucose uptake, and this could be confirmed in human systems in the future.

## Data Availability

All data produced in the present work are contained in the manuscript and in the PRIDE proteomeXchange repository with accession number PXD032948.

https://www.ebi.ac.uk/pride/

## ACKNOWLEDGEMENTS

We thank Betina Bolmgreen, Nikoline R. Andersen, Jesper B. Birk and Irene B. Nielsen (Department of Nutrition, Exercise and Sports) for their skilled technical help in the laboratory. We would like to thank Professor Bente Kiens for scientific discussion, Kasper E. Villumsen and Kim A. Sjøberg for assistance during the experimental trials and Le Lene S. S. Stevner for support on data regulation and protection. This work was funded by a Pfizer, Inc. grant (to D.E.J., J.F.P.W., C.P.); and a grant from the National Health and Medical Research Council (NHMRC; GNT2011083; to S.J.H., D.E.J., J.F.P.W.). E.J.N. was supported by an Australian Government Research Training Program Scholarship, the University of Sydney Val Street Scholarship, and was supported by the Schmidt Science Fellows, in partnership with the Rhodes Trust; S.J.H. is supported by an NHMRC Investigator Grant (2026905); D.E.J. is an Australian Research Council (ARC) Laureate Fellow; The Novo Nordisk Foundation (no. NN0070370, NNF082659 and NNF0079480 to J.F.P.W.); The University of Sydney University of Copenhagen Partnership Collaboration Awards (to J.F.P.W. and D.E.J.); J.D.O. and M.R.L. were supported by a research grant from the Danish Diabetes and Endocrinology Academy, which is funded by the Novo Nordisk Foundation, grant number NNF17SA0031406. Japan Society for the Promotion of Science Grants-in-Aid for Scientific Research 19K20007 (to K.K.), and the European Foundation for the Study of Diabetes JDS Fellowship Program (to K.K.). B.L.P. is supported by an NHMRC Investigator Grant (2009642).

E.J.N was supported by core funding from EMBO (ALTF 1071-2021), the: British Heart Foundation (RG/18/13/33946), NIHR Cambridge Biomedical Research Centre (BRC-1215-20014; NIHR203312), Cambridge BHF Centre of Research Excellence (RE/18/1/34212), BHF Chair Award (CH/12/2/29428) and by Health Data Research UK, which is funded by the UK Medical Research Council, Engineering and Physical Sciences Research Council, Economic and Social Research Council, Department of Health and Social Care (England), Chief Scientist Office of the Scottish Government Health and Social Care Directorates, Health and Social Care Research and Development Division (Welsh Government), Public Health Agency (Northern Ireland), British Heart Foundation and Wellcome. The content is solely the responsibility of the authors and does not necessarily represent the official views of the NHMRC, ARC, the NIHR or the Department of Health and Social Care. The authors acknowledge the facilities and support of the Sydney Mass Spectrometry Facility (SydneyMS), University of Sydney.

## AUTHOR CONTRIBUTIONS

Conceptualization, C.P, D.E.J, J.F.P.W, S.J.H, B.L.P, E.J.N, J.R.H; Clinical screening, J.R.H, J.M.K, K.K, K.H, J.D.O; Formal analysis, E.J.N, S.J.H, J.R.H, J.D.O, A.D.V, M.R.L; Investigation E.J.N, J.R.H, S.J.H, J.D.O, A.D.V, M.R.L, J.M.K, EAR, K.K, K.C, G.Y, J.F.P.W; Resources, J.F.P.W, D.E.J, S.J.H; Data Curation, S.J.H, E.J.N, J.R.H, J.D.O; Writing - Original Draft, E.J.N, D.E.J, S.J.H, J.F.P.W; Writing – Review and Editing *all authors*; Visualization, E.J.N, S.J.H; Supervision, D.E.J, S.J.H, J.F.P.W; Funding Acquisition C.P, J.F.P.W, D.E.J.

## SUPPLEMENTAL INFORMATION

**Table S1:**
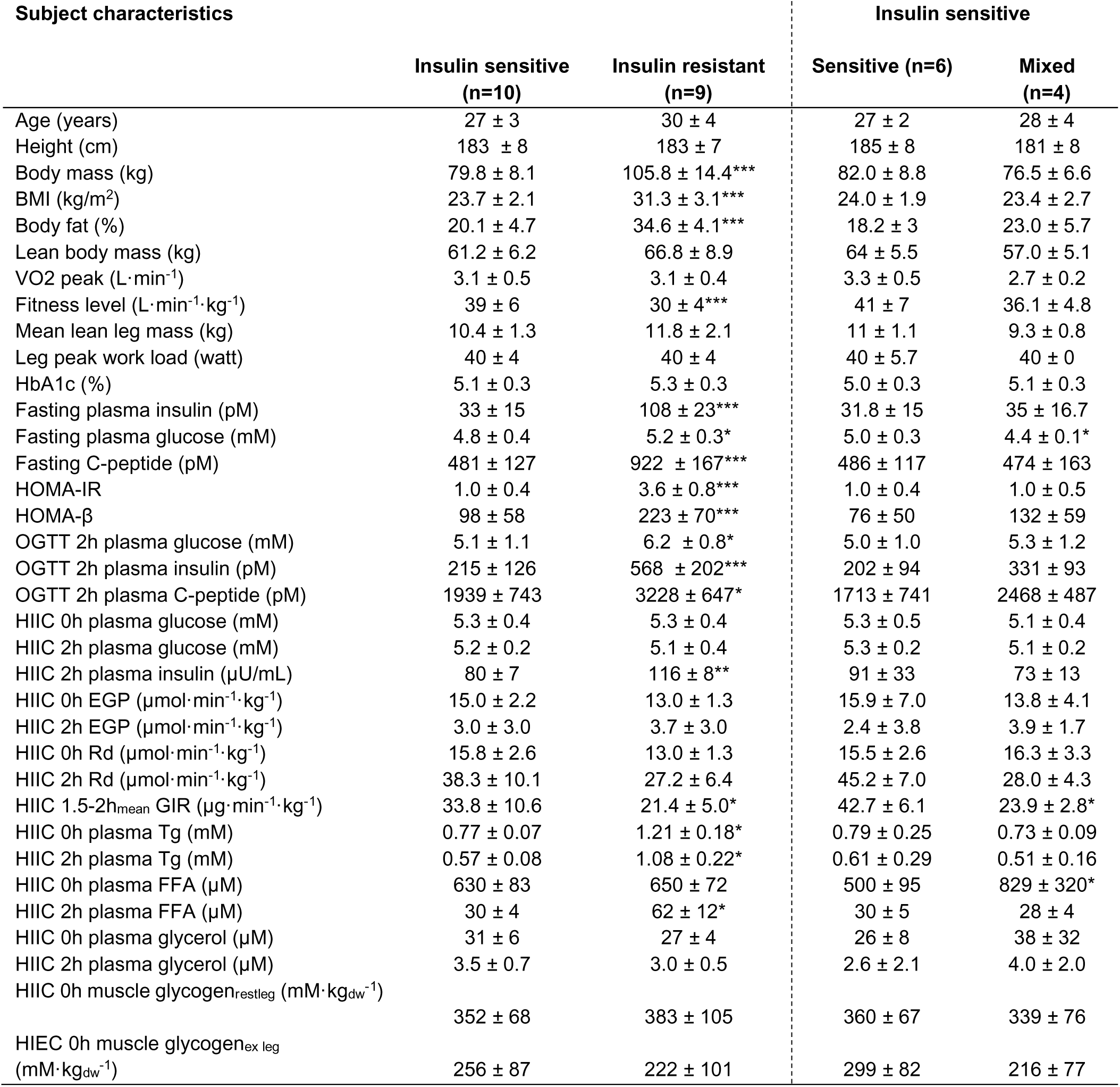
Subject characteristics of both insulin sensitive and insulin resistant subjects. Insulin sensitive group is subdivided into a mixed and a sensitive subgroup based on glucose infusion rate during the hyperinsulinemic isoglycemic clamp as described in the main text. HOMA-IR calculated as Plasma Glucose (mM) · Plasma Insulin (µU/mL) ·22.5^−1^. HOMA-beta calculated as (20 · Plasma Insulin (µU/mL)) · (Plasma Glucose (mM) – 3.5)^−1^. Data presented as means ± SD. *P<0.05, **P<0.01, ***P<0.001 difference between Insulin Sensitive group and Insulin resistant Group or Sensitive or Mixed subgroups.

**Figure S1:**
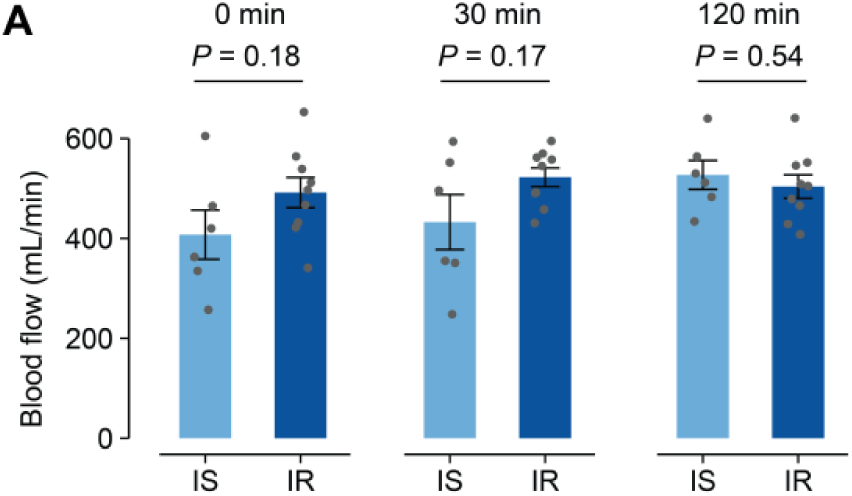
Bulk blood flow does not differ between insulin sensitive and insulin resistant subjects. **(A)** Blood flow of the non-exercised leg during a hyperinsulinemic-euglycemic clamp. Error bars represent mean +/- SEM.

**Figure S2:**
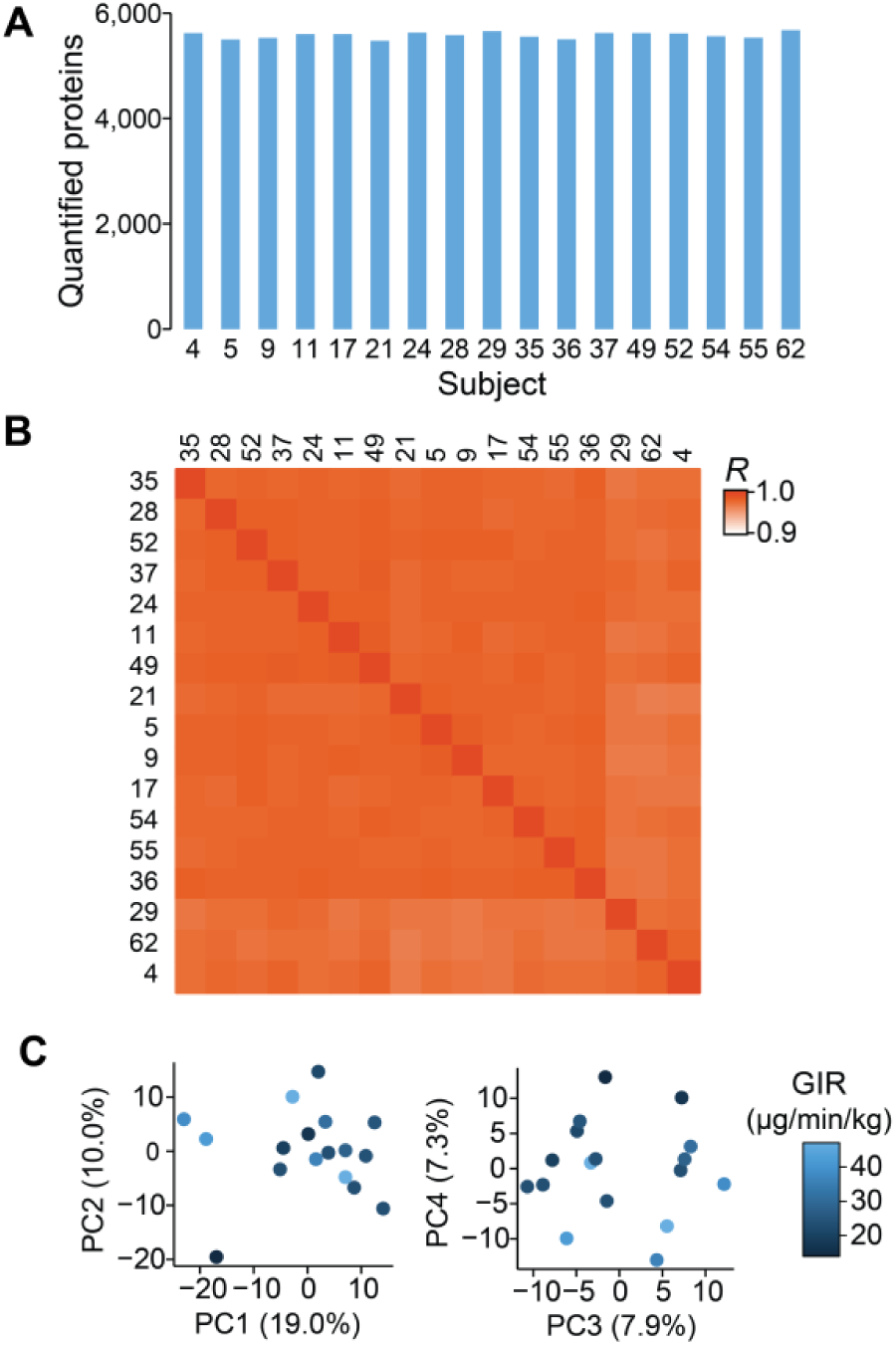
Proteome quality control. (**A**) Number of quantified proteins per sample. (**B**) Correlation matrix of the proteomes. (**C**) Principal component analysis of the proteomes coloured by steady-state GIR during an insulin clamp.

**Figure S3:**
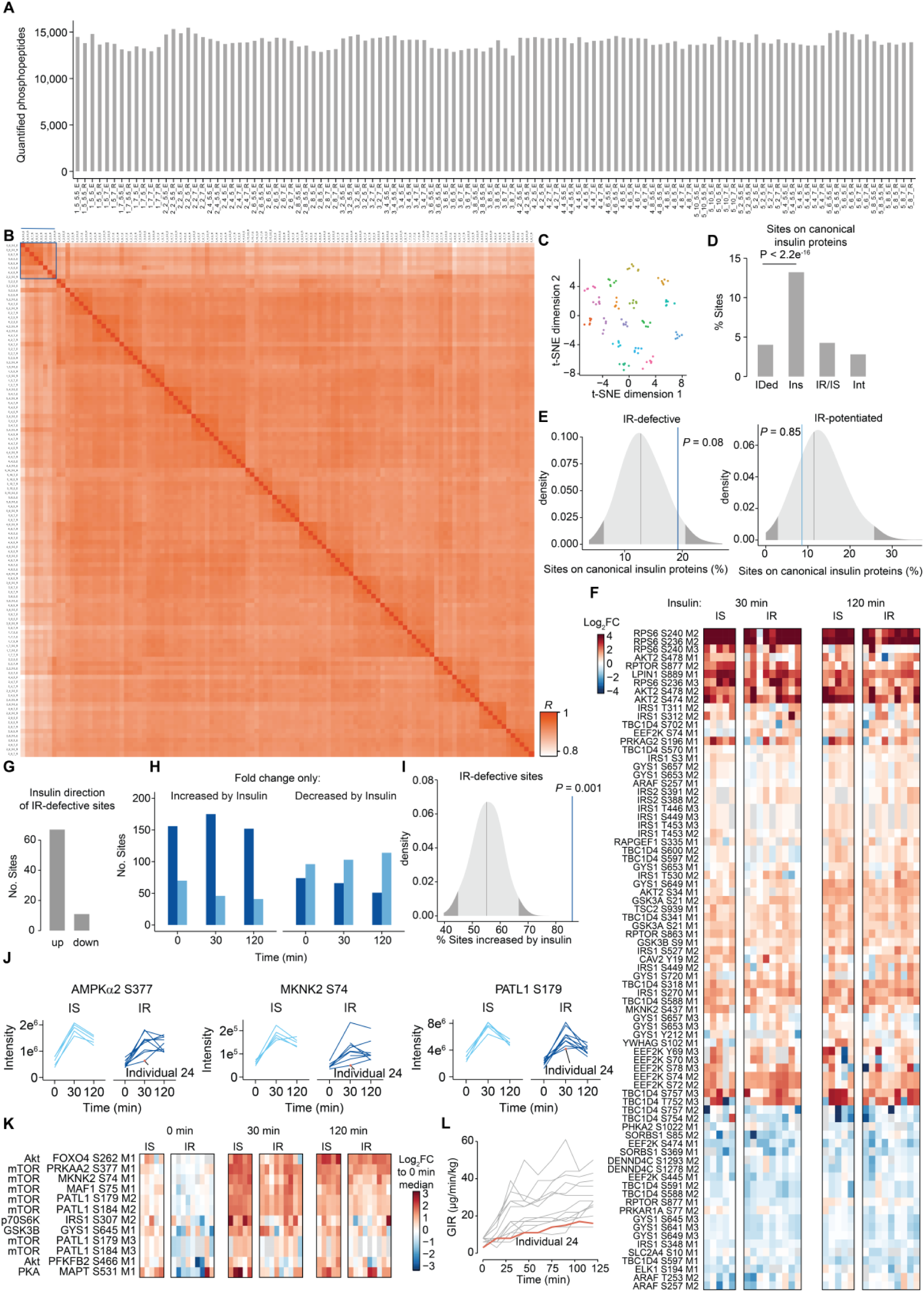
Defective phosphosites in insulin resistance. **(A)** The number of quantified phosphopeptides per sample. **(B)** Correlation matrix of the phosphoproteomes. A line is drawn above the samples that were identified to be outliers and are suspected to include higher levels of non-muscle cell-types. **(C)** Clustering by t-SNE of the phosphoproteomes of all biopsies, coloured by individual. **(D)** The percentage of phosphosites that occur on proteins involved in canonical insulin signalling out of all the quantified phosphosites, and the regulated phosphosites in each condition (*P*_adj_ < 0.05 and FC > 1.25) (Fisher’s exact test). **(F)** Heatmap of the phosphosites on canonical insulin signalling proteins that are equivalent between insulin sensitive and insulin resistant individuals for potentiated in IR sites. **(G)** The direction that insulin regulates the phosphosites with defective insulin regulation in IR individuals. **(H)** The number of phosphosites that are regulated by insulin and changed by at least 1.25-fold between IR vs IS individuals, coloured by whether IR is less than the direction of insulin regulation in IS subjects (defective) or greater than (potentiated). **(I)** Permutations of the percentage of sites occurring on canonical insulin signalling proteins compared to those that are defective and potentiated in insulin resistant individuals. **(J)** Example profiles of reported mTOR substrates that are defective in insulin resistant individuals, with the outlier individual no.24 labelled. **(K)** Heatmap of phosphosites with defective insulin regulation in IR individuals and a reported upstream kinase. **(L)** GIR during a hyperinsulinemic-euglycemic clamp with individual 24 coloured in orange.

**Figure S4:**
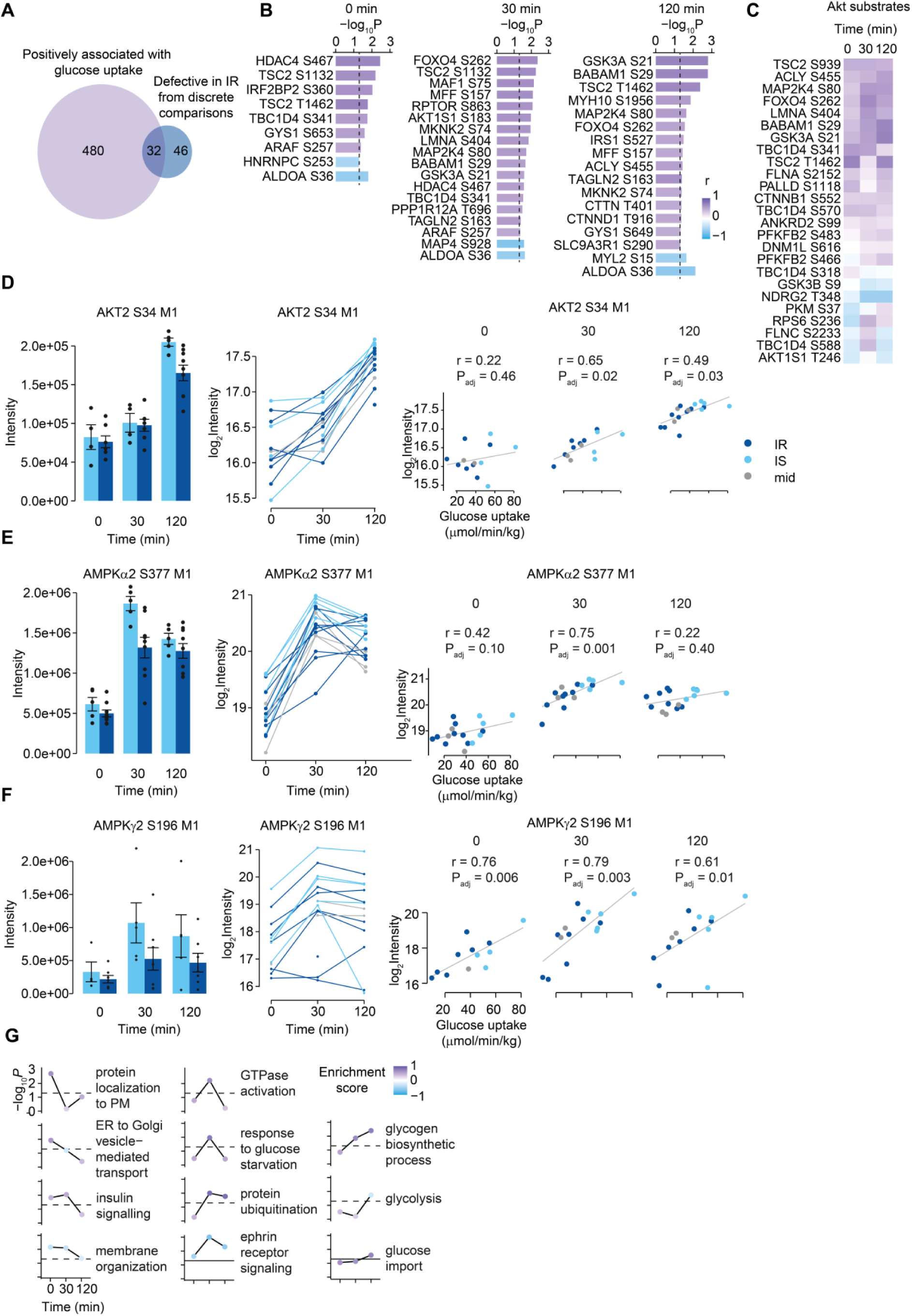
Phosphosites associated with the degree of insulin sensitivity across individuals. (**A**) Venn diagram of the common steady state glucose uptake-associated phosphosites (*P*_adj_ < 0.05) and the defective phosphosites in IR individuals from discrete comparisons (*P*_adj_ < 0.05, FC > 1.25). (**B**) Phosphosites with a reported function (PhosphosSitePlus) that are associated with rested-leg steady state glucose uptake. (*P*_adj_ < 0.05). (**C**) Heatmap of correlation-coefficients of AKT substrates with glucose uptake. The levels of (**D**) AKT2 S34, (**E**) AMPKa2 S377 and (**F**) AMPKg2 S196 in IR (navy) and IS (light blue) individuals at the different timepoints of the hyperinsulinemic-euglycemic clamp, as a bar plot and repeated measures profile, and their correlations with steady-state glucose uptake. (**G**) Enrichment of GO biological processes in the groups of proteins with phosphosites measured at the different timepoints associated with LGU.

**Figure S5:**
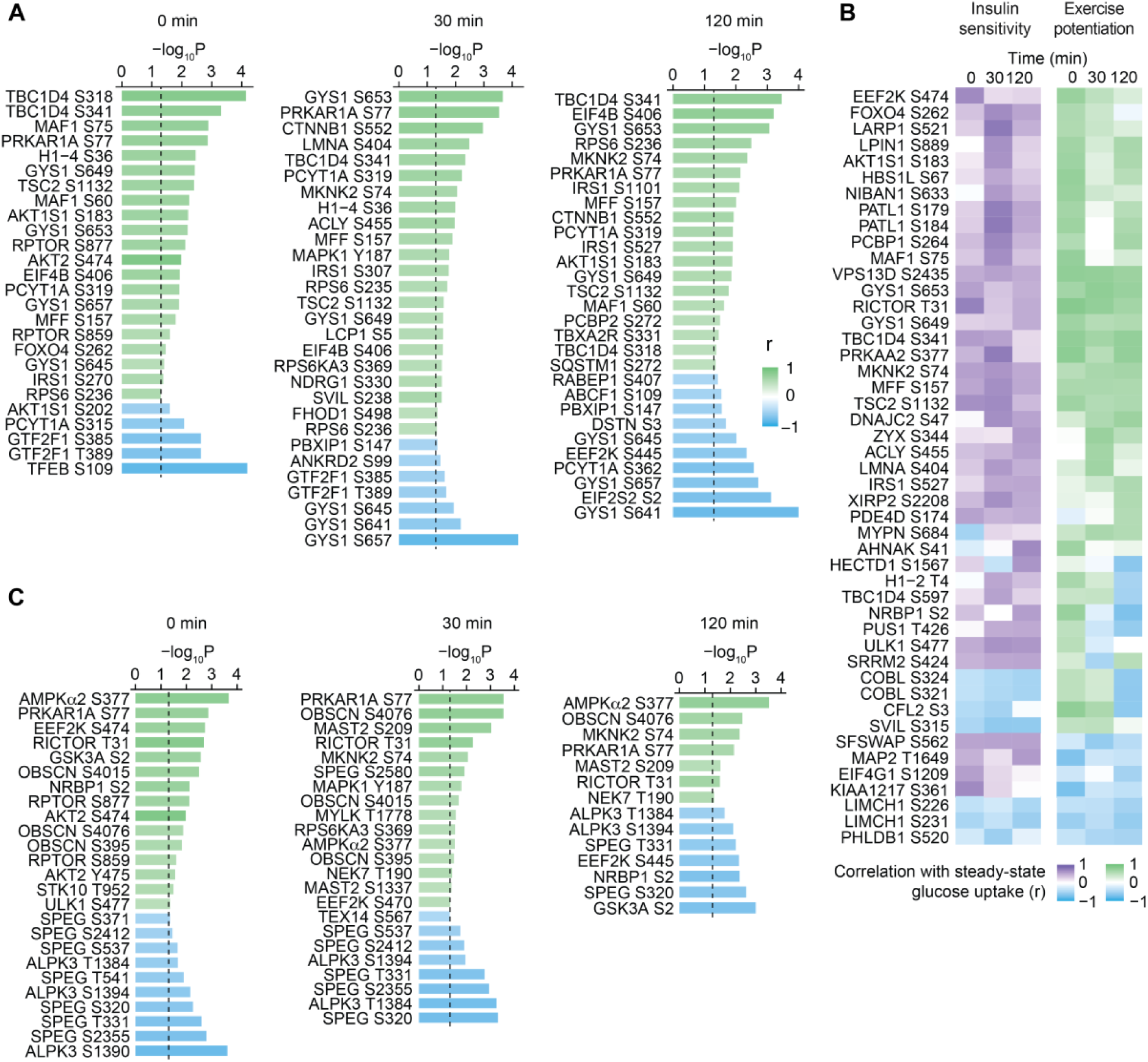
Phosphosites associated with the potentiation of steady state glucose uptake by exercise. **(A)** Phosphosites with a reported function (PhosphosSitePlus) that are associated with steady state glucose uptake across the exercised and non-exercised leg (*P*_adj_ < 0.05). **(B)** Phosphosites that are associated with both LGU in the non-exercised leg and exercise-mediated potentiation LGU at any timepoint (*P*_adj_ < 0.05). **(C)** Phosphosites on kinases that are associated with the potentiation of steady state glucose uptake by exercise (*P*_adj_ < 0.05).

**Figure S6:**
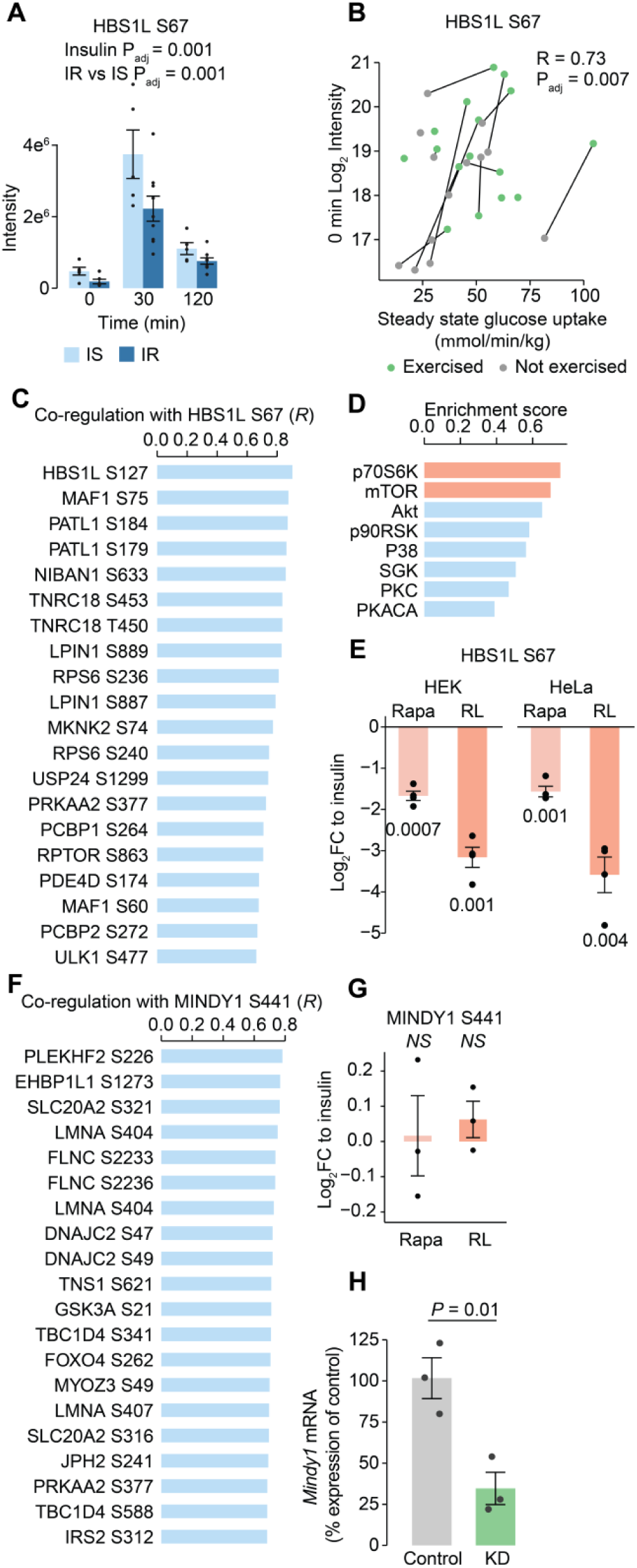
Utilising the co-regulation of phosphosites to predict upstream kinases of uncharacterised sites. **(A)** Intensity of HBS1L S67 in the non-exercised leg, comparing IS and IR groups. **(B)** Repeated measures correlation of HBS1L S67 in the non-exercised and exercised leg at 0 min with the LGU. **(C)** The 20 most highly co-regulated phosphosites with HBS1L S67. **(D)** Enrichment of kinase substrates in all sites ordered by their co-regulation with HBS1L S67. **(E)** HBS1L S67 abundance after treatment of HEK and HeLa cells with 100 nM insulin and either 100 nM rapamycin (rapa) or rapalink (RL), which inhibit mTORC1. RL was used at 1 nM in HeLa cells and 3 nM in HEK cells. Both cell lines were treated with 100 nM rapamycin. **(F)** The 20 most highly co-regulated phosphosites with MINDY1 S441. **(G)** MINDY1 S441 levels after treatment of HEK cells with insulin and 100 nM rapamycin or 3 nM rapalink. MINDY1 S441 was not quantified in HeLa cells. **(H)** *Mindy1* mRNA levels after knockdown of *Mindy1* compared to a non-targeting siRNA control (N = 3 biologically independent samples). Error bars represent mean +/- SEM.

## METHODS

### Human physiology

This study was approved by the Committee on Health Research Ethics of the Capital Region of Denmark (Journal number: H-18017861) and performed in accordance with the standards set by the Declaration of Helsinki II. All subjects gave written informed consent. The study was registered at Clinical Trial # NCT04178603.

Male subjects were screened for normal haematology parameters, no family history of diabetes and a normal glucose tolerance (plasma glucose <7.8 mmol l^−1^ in a 2-h oral glucose tolerance test). The subjects were subdivided into two groups fulfilling the criteria of insulin sensitive and normal weight (HOMA-IR < 1.5, BMI < 25 kg/m^2^, n = 10) or insulin resistance and overweight (HOMA-IR >= 2.2, BMI = 28-35 kg/m^2^, n = 9). Body composition was determined by DXA (DPX-IQ Lunar; Lunar Corporation, Madison, WI). The peak oxygen uptake was measured (MasterScreen CPX; Intramedic A/S, Gentofte, Denmark) during cycling exercise (Monark, Vansbro, Sweden) during an incremental exercise test. Furthermore, the study participants were familiarized to the one-legged knee extensor ergometer on several occasions, and at a minimum of 1 week prior to the first experimental day, peak workload (PWL) of the knee extensors was determined in both legs by an incremental test^82^.

After refraining from alcohol, tobacco, nicotine and exercise for 48 h, subjects met at 07:00 in the morning after an overnight fast. Following arrival at the laboratory, subjects ingested a light, standardized breakfast (oatmeal, skimmed milk and sugar— 5% of daily energy intake). Subjects then rested until exercise was initiated at 09:00. The exercise protocol consisted of one-legged knee-extensor exercise for 1 h at 80% PWL, interspersed with three 5-min intervals of 100% PWL (last 5 min of each 20 min periode). Upon conclusion of exercise, subjects returned to a supine, resting position, where catheters (Pediatric Jugular Catheterization Set, Arrow International, Reading, PA) were inserted into a femoral artery and both femoral veins during local anesthesia (10 mg/mL lidocaine without adrenaline, Xylocaine; AstraZeneca, London, UK). After 2 hours of rest, infusion of a sterile uniform 13C-glucose tracer (U-13C6 D-glucose, Cambridge Isotope Laboratories, Inc., Massachusetts, USA) was initiated to reach and maintain a 10% blood glucose enrichment. After 2 hours of tracer infusion a 2-hour hyperinsulinemic euglycemic clamp was performed with a bolus of insulin (9mU·kg^−1^) (Actrapid; Novo Nordisk, Bagsværd, Denmark) followed by a constant insulin infusion rate (1.42 mU·kg^−1^·min^−1^). Throughout the clamp, 20% glucose enriched with 10% ^13^C-glucose tracer was infused with varying rates to secure euglycemia and blood glucose tracer enrichment. To measure insulin-stimulated leg glucose uptake in both legs before and during the clamp, the A-V balance technique was combined with ultrasound Doppler measurements (Philips iU22; ViCare Medical A/S, Birkerød, Denmark) of femoral arterial blood flow. Muscle biopsies from m. vastus lateralis were collected at the start, 30 minutes into and at the end of the clamp (120 min) from both the prior exercised and rested leg, rinsed in ice-cold saline, and snap-frozen in liquid nitrogen.

The 0 and 120 minute biopsies from the lean insulin sensitive subjects from the study (HOMA-IR < 1.5, BMI < 25 kg/m^2^, n = 10) were previously published as a validation cohort^19^. Blood gases and plasma glucose and lactate were measured (ABL800 FLEX, Radiometer, Denmark) concurrent with blood flow measurements (Before, and every 15 min during the clamp). Serum insulin and C-peptide levels were analysed on Cobas e411 (Roche Diagnostics International Ltd., California).

### Sample preparation

#### Skeletal muscle phosphoproteomes

Frozen muscle biopsies (40-60 mg wet weight) were ground on liquid nitrogen with a mortar and pestle, lysed in 175 µL 6 M GdmCl/100 mM Tris pH 8.5 and sonicated (5 min., 90% amplitude, 2s/5s pulses, 4 °C) in a Q800R2 (QSonica). Lysates were immediately boiled (95 °C 5 min.), then sonicated (1 min., 90% amplitude, 5s/5s pulses, 4 °C), and centrifuged at 20,000 xg for 5 min. Supernatants were reduced/alkylated with 10 mM TCEP/40 mM CAA (5 min., 45 °C followed by 40 min., RT). Protein was precipitated with chloroform:methanol (1.25 volumes of 1:4), and resolubilised with 6 M GdmCl/100 mM Tris pH 8.5 by sonication. Samples were diluted 1:1 with water and protein precipitated with 4 volumes of acetone (-30 °C overnight). Precipitated protein was resuspended in SDC buffer (4% SDC/100 mM Tris pH 8.5) with sonication, 1.2 mg of protein was processed with the EasyPhos workflow^25^, and 25 µg of non-phosphorylated peptides (EasyPhos supernatant) was retained for proteome analysis.

#### Skeletal muscle proteomes

25 µg of non-phosphorylated peptides from each muscle sample was retained as the flowthrough from phosphopeptide enrichment and subjected to fractionation for proteome analysis. Peptides were resuspended in 100 µl of MS loading buffer (2% ACN 0.3% TFA) and fractionated using a Dionex UltiMate 3000 HPLC, on an XBridge Peptide BEH C18 column (130 Å, 3.5 μm, 2.1 mm x 250 mm), maintained at 50°C using the U3000 column oven. Mobile phases comprised Buffer A (10 mM ammonium formate/2% ACN) and Buffer B (10 mM ammonium formate/80% ACN) adjusted to pH 9.0 with NH_4_OH. Peptides were separated with a gradient of 10 – 40% buffer B over 4.4 mins, followed by 40 – 100% buffer B over 1 min. 72 fractions were collected with concatenation during collection into 24 samples so that fractions #1, #25 and #49 were pooled (as were fractions #2, #26 and #50, etc.). After fraction collection samples were dried down directly in the deep-well plate using a vacuum concentrator (GeneVac).

### Liquid chromatography-tandem mass spectrometry (LC-MS/MS)

#### Skeletal muscle phosphoproteomes

Enriched phosphopeptides in MS loading buffer (2% ACN, 0.3% TFA) were loaded onto in-house fabricated 50-cm columns with a 75-µM I.D., packed with 1.9 µM C18 ReproSil Pur AQ particles using a Dionex U3000 HPLC coupled to an Orbitrap Exploris 480 mass spectrometer (Thermo Fisher Scientific). Column temperature was maintained at 60°C with a Sonation column oven, and peptides separated using a binary buffer system comprising 0.1% formic acid (buffer A) and 80% ACN plus 0.1% formic (buffer B), at a flow rate of 400 nl/min, with a gradient of 3–19% buffer B over 80 min followed by 19–41% buffer B over 40 min, resulting in ∼2-h gradients. Peptides were analysed with one full scan (350–1,400 m/z, R = 120,000) at a target of 3e6 ions, followed by 48 data-independent acquisition (DIA) MS/MS scans (350–1,022 m/z) with higher-energy collisional dissociation (HCD) (target 3e^6^ ions, max injection time 22 ms, isolation window 14 m/z, 1 m/z window overlap, normalised collision energy (NCE) 25%), with fragments detected in the Orbitrap (R = 15,000).

#### Skeletal muscle proteomes

Fractionated proteome peptides in MS loading buffer (2% ACN 0.3% TFA) were loaded onto a 150 µm ID x 15 cm column packed in-house with 1.9 μm C18 material (ReproSil Pur AQ C18, Dr. Maisch, GmbH) using an EvoSep One HPLC. Column temperature was maintained at 60°C using a column oven (Sonation). Peptides were separated using a binary buffer system comprising 0.1% formic acid (buffer A) and ACN plus 0.1% formic acid (buffer B) using the standardized “30 SPD” (Samples Per Day) method, resulting in MS acquisition times of 44 minutes plus overhead of ∼2 minutes per sample. Peptides were directly sprayed into an Orbitrap Exploris 480 mass spectrometer (Thermo Fisher Scientific) with a spray voltage of 2.4 kV, and analysed in Data Independent Acquisition (DIA) format, with one full scan (350–1,400 m/z, R = 120,000) at a target of 3e6 ions, followed by 48 DIA MS/MS scans (350– 1,022 m/z) with higher energy collisional dissociation (HCD) (target 3e6 ions, max injection time 22 ms, isolation window 14 m/z, 1 m/z window overlap, normalised collision energy (NCE) 25%), with fragments detected in the Orbitrap (R = 15,000).

### RAW data processing

RAW data was analysed using Spectronaut (v14.11.210528.47784). Data were searched using directDIA against the Human UniProt Reference Proteome database (December 2019 release), with default settings (precursor and protein Qvalue cutoffs 0.01, Qvalue filtering, MS2 quantification), with “PTM localization” turned on. The PTM “Probability cutoff” was set to 0 and localization filtering (min. 0.75 localization probability) was performed during downstream analysis. Spectronaut output tables were processed using the Peptide collapse (v1.4.2) plugin for Perseus^83,84^.

### Quality control and processing

#### Phosphoproteomics

Intensity values were log_2_ transformed and median normalised. For quality control, a correlation matrix of Pearson’s correlation for all pairwise observations was generated. 8 samples out of 114 (“1_5_5_E”, “2_2_5_R”, “2_2_5.5_R”, “5_6_5_E”, “5_6_5_R”, “5_6_5.5_E”, “5_6_5.5_R”, “5_6_7_R”) were outliers. These samples had high quality enrichments and MS runs but we hypothesised that these biopsies may be contaminated by non-muscle tissue as they contained lower phosphorylation levels of muscle markers and were excluded from subsequent analysis.

#### Proteomics

To explore ground-state differences between insulin resistant and insulin sensitive skeletal muscle, the proteomes of the biopsies at time 0 of the insulin clamp in the non-exercised leg were measured. Since the samples “2_2_5_R” and “5_6_5_R” were outliers suspected to contain non-muscle tissue based on phosphoproteomes, we did not measure proteomes of these samples.

### Proteomics data analysis

#### Statistical testing

To compare insulin sensitive and insulin resistant proteomes, t-tests were performed with empirical bayes moderation of standard error using LIMMA^85^. The “mixed” samples were not included in this test. Empirical *P*-values were calculated by permuting the subject labels 1,000 times.

### Phosphoproteomics data analysis

#### Statistical testing

Two multilevel linear models were performed. The first tested the difference between IR and IS individuals and included the factors of insulin as a within-subject factor and IR vs IS between subjects, and their interaction. P-values < 0.05 were filtered for multiple hypothesis testing correction by permuting the labels 1,000 times to calculate empirical *p*-values. The second tested the effect of exercise on insulin signalling within subjects, and so included the factors of insulin and exercise as within-subject factors and their interaction. Due to the greater power of the complete within-subject comparison, a more stringent P-value adjustment method was used than permutations. P-values were adjusted to q-values with the R package qvalue^86^. This method is a more powerful version of BH P-value correction as is estimates the proportion of true null hypotheses rather than assuming all hypotheses are null as in BH.

#### Personalized phosphoproteomics associations

##### Phosphosites associated with insulin sensitivity in the non-exercised leg

“Steady state” glucose uptake was calculated as the mean of the 90, 105 and 120-minute leg glucose uptake measures in the non-exercised leg during the clamp. Phosphosite abundances from 0, 30 and 120-minute timepoints were separately correlated with steady state glucose uptake for a temporally shifted application of personalized phosphoproteomics. Steady-state glucose uptake values were normally distributed (Shapiro-Wilk test *P* > 0.05), so Pearson’s correlations were performed. Phosphosites were tested for association if they were a) quantified in at least 10 subjects at that timepoint b) significantly regulated, determined by a significant main effect or interaction in the multilevel linear model testing the effects of insulin and IR vs IS (*P_adj_* < 0.05, FC > 1.25), c) Had a sufficient differences across subjects at that timepoint (>= 1.5-fold between the maximum and minimum). Duplicate phosphopeptides with identical quantification due to multiple phosphosites being present were collapsed to single values. For sites passing non-adjusted *P* < 0.05, empirical P-values were calculated by permuting the glucose uptake subject labels 1000 times then performing the associations.

*Phosphosites associated with exercise potentiating insulin-stimulated glucose uptake:* Steady state glucose uptake was calculated as above for both the exercised and non-exercised legs. Phosphosites were filtered as above, except for regulation in the exercise linear model rather than the linear model testing IR vs IS. To associate the effect of prior exercise within each subject on their steady state glucose uptake, repeated measures correlations were performed to incorporate the non-exercised and exercised leg values for each timepoint separately, utilising the rmcorr R package^52^. Empirical P-values were calculated as above.

### Databases

Kinase substrate relationships were obtained from PhosphoSitePlus Kinase Substrate Dataset (January 2024 version) and substrates of kinase isoforms, for example AKT1 and AKT2, were merged to broader groupings^87^. Regulatory sites were obtained from the PhosphoSitePlus database of the same name (January 2024 version). Pathways were GO biological processes. Nearest genes to published genetic variants associated with type 2 diabetes, fasting glucose, HbA1c, fasting insulin or 2hr glucose during an oral glucose tolerance test were obtained from previously published studies^50,62^. The T2D knowledge portal was accessed on 12 February 2024 for information about MINDY1 genetic variants.

### Enrichment analyses

Overrepresentation of kinase substrates in specified groups of phosphosites were tested with one-sided Fisher’s Exact tests, using all the quantified phosphosites in this study as background. *P*-values were adjusted with the q-value method. Kinase or pathway enrichments were performed on sites ordered by either fold change or correlation co-efficient with a modified two-sided Kolmogorov-Smirnov test implemented in the R package ksea, which calculates empirical P-values by 1,000 permutations^88^.

### mTOR inhibitor studies

Rapamycin and Rapalink phosphoproteome studies were performed as described^19^. Briefly, SILAC-labelled HEK293E cells were serum-starved for 4 hours together with either 100 nM rapamycin, 3 nM or 10 nM RapaLink1, or vehicle (DMSO), and treated with 100 nM insulin or vehicle (10 min., 37°C), scraped in ice-cold lysis buffer (6M GdmCl, 100 mM Tris pH 8.8, 10 nM TCEP, 40 mM CAA), and processed with the EasyPhos method^89^. Experiments were performed with four biological replicates.

### Glucose uptake experiments

#### Cell culture

Experiments were conducted using Mycoplasma-free L6 myotubes overexpressing HA-GLUT4 cell lines. HA-GLUT4 overexpression is crucial for investigating insulin sensitivity in vitro, as previously outlined^90^. L6-myoblasts were cultured in Dulbecco’s Modified Eagle Medium (DMEM) supplemented with 10% fetal bovine serum (FBS) and 2 mM GlutaMAX at 37°C with 10% CO2. The myoblasts were differentiated in DMEM/GlutaMAX containing 2% horse serum, with media replaced every 48 hours for 6 days. Experiments were conducted using L6 myotubes on day 7 post-differentiation, ensuring at least 90% cell differentiation prior to experimentation.

#### Protein knockdown by siRNA transfection

Protein knockdown using siRNA was conducted in L6-myotubes on day 4 post-differentiation. Briefly, Opti-MEM (Thermo Fisher Scientific) and TransIT-X2 (Mirus, MIR6006) were mixed at a ratio of 30:1 and incubated at room temperature for 20 minutes. siRNA was then added to the Opti-MEM/TransIT-X2 mixture to achieve a final concentration of 50 nM per well siRNA (ON-TARGETplus SMART POOL, 4-siRNA/target), gently mixed by pipetting, and incubated at room temperature for 30 minutes. Mindy1 SMART POOL sequences were: 1. UGGACGUCAAUGUGCGAUU; 2. GGAUAACCUGUAUGCGCUA, 3. AGUUAUAACCAGCUAGUAG and 4. AGGUAAAGCUGCCGCCUCA. Non-targeting SMART POOL sequences were: 1. UGGUUUACAUGUCGACUAA, 2. UGGUUUACAUGUUGUGUGA, 3. UGGUUUACAUGUUUUCUGA, and 4. UGGUUUACAUGUUUUCCUA. The transfection reagents were subsequently transferred into cells seeded in 24-well plates (for glucose uptake and RNA extraction). Media were replaced 24 hours after transfection with DMEM supplemented with 2% HS and GlutaMAX. Insulin-induced glucose uptake was performed day 7 post-differentiation.

#### Glucose uptake

Following 16 hours of serum starvation, we assessed glucose transporter activity using established protocols^91^. Briefly, cells were washed three times with PBS after treatment and then incubated with Krebs-ringer 0.2% BSA buffer (KRP buffer; composed of 0.6 mM Na2HPO4, 0.4 mM NaH2PO4, 120 mM NaCl, 6 mM KCl, 1 mM CaCl2, 1.2 mM MgSO4, and 12.5 mM Hepes [pH 7.4]). Subsequently, cells were stimulated with 100 nM insulin for 20 minutes. To assess non-specific glucose incorporation, 25 μM of cytochalasin B (dissolved in ethanol; Sigma-Aldrich) was added before the introduction of 2-[3H]deoxyglucose (2-DOG) (PerkinElmer). During the final 5 minutes of stimulation, 2-DOG (0.25 μCi, 50 μM) was introduced. Afterward, cells were washed three times with ice-cold PBS, and lysate with 1 mM NaOH followed by neutralisation with 1 mM of HCl, and 2-DOG levels were determined using liquid scintillation counting. The obtained data were normalized based on protein concentration and further adjusted relative to basal control cells.

#### RT-PCR

Cells were subjected to total RNA extraction using TRIzol reagent (Invitrogen), following the manufacturer’s instructions, except for using 1-bromo-3-chloropane (Sigma) instead of chloroform for phase separation. RNA precipitation was performed using isopropanol, followed by washing with 70% ethanol and reconstitution in DEPC water. The quality and quantity of RNA were assessed using a Nanodrop 2000 spectrophotometer. Subsequently, 500 ng of RNA was reverse transcribed into cDNA using PrimeScript Reverse Transcriptase (Clontech, Takara Bio Company) according to the manufacturer’s instructions with oligo-dT primers. PCR reactions were conducted using 1:10 diluted cDNA and SYBR Select Master Mix (Thermo Fisher Scientific) on the LightCycler 480 II (Roche), employing the following primer sets: Mindy1 F 5′ GACCAGGACTACCTTATTGCCT 3′ and Mindy1 R 5′ AGGTCACTGAGACCCAGCAT 3′. All samples were normalized to the housekeeping gene Beta-actin using the primer pair B-actin F 5′ GAGATTACTGCCCTGGCTCCTA 3′ and B-actin R 5′ GACTCATCGTACTCCTGCTTGCTG 3′. The efficiency of the assays was verified using serial dilutions of a pooled control sample, and only primers exhibiting efficiencies within the range of 85–120% were utilised.

